# The yield of tuberculosis contact investigation in São Paulo, Brazil: a community-based cross-sectional study

**DOI:** 10.1101/2024.06.26.24309533

**Authors:** José Mário Nunes da Silva, Fredi Alexander Diaz-Quijano

## Abstract

The strategy of tuberculosis (TB) contact investigation is essential for enhancing disease detection. We conducted a cross-sectional study to evaluate the yield of contact investigation for new TB cases, estimate the prevalence of TB, and identify characteristics of index cases associated with infection among contacts of new cases notified between 2010 and 2020 in São Paulo, Brazil. Out of 186,466 index TB cases, 131,055 (70.3%) underwent contact investigation. A total of 652,286 contacts were screened, of which 451,704 (69.2%) were examined. Of these, 12,243 were diagnosed with active TB (yield of 2.0%), resulting in a number needed to screen of 51 and a number needed to test of 37 to identify one new TB case. The weighted prevalence for the total contacts screened was 2.8% (95% confidence interval [CI]: 2.7%–2.9%), suggesting underreporting of 6,021 (95% CI: 5,269–6,673) cases. The likelihood of TB diagnosis was higher among contacts of cases with active case-finding, abnormal chest X-ray, pulmonary TB, or drug resistance, as well as among children, adults, women, individuals in socially vulnerable situations, and those with underlying clinical conditions. The study highlights significant TB underreporting among contacts, recommending strengthened contact investigation to promptly identify and treat new cases.

## Introduction

Tuberculosis (TB) remains the second leading cause of death from infectious diseases worldwide, posing a significant public health concern [1]. In 2022, there was a notable 28% increase in the global number of newly diagnosed cases compared to 2020 [1]. Nevertheless, the global targets set in 2018 regarding treatment, prevention, and funding have not been met, and efforts to reduce this burden remain insufficient [2]. To reverse this trend, it is crucial for each country to intensify the identification and proper treatment of TB cases, aiming to achieve the global goal of ending the epidemic by 2035 [1,3].

TB contact investigation is a crucial and cost-effective strategy [4], aimed at enhancing disease detection [5] and improving treatment outcome [6]. Its primary goal is to promptly identify and treat any secondary cases of the disease, as well as to detect contacts with latent TB infection (LTBI) eligible for preventive treatment [7]. Additionally, it plays a pivotal role in tracing the source case, particularly for children under 5 years old diagnosed with TB, facilitating the implementation of appropriate control measures [7]. The effectiveness of the investigation is assessed by its yield, which is the percentage of screened contacts found to have TB [3].

In a meta-analysis of 181 studies, the combined global prevalence of TB from contact investigations was 3.6%. This proportion varied from 5.0% in low-income countries to 4.4% in middle-income countries, while in high-income countries it was 1.8% [8]. In Brazil, a country with high middle-income status and a high TB burden, the incidence of TB among household contacts is estimated at 427.8 per 100,000 person-years at risk, approximately 16 times the incidence in the general population [9], with prevalence potentially reaching 5.7% [10]. Since 2009, it has been recommended that all close contacts of a smear-positive pulmonary TB case, regardless of symptoms, age, and HIV status, undergo investigation for active TB or LTBI [11].

However, in 2023, only 53.9% of the identified contacts of laboratory-confirmed new pulmonary TB cases were examined [12]. Moreover, there is a lack of comprehensive information on the yield of this strategy in routine programmatic settings, as well as whether individual characteristics of index cases are associated with a higher likelihood of TB infection among contacts.

Understanding these factors can assist national programmes adapt their contact investigation strategies to improve their effectiveness and yield, especially in high-incidence settings [8,13]. Therefore, our aims were to evaluate the yield of community-based contact investigations for new TB cases, estimate disease prevalence among contacts, and identify which characteristics of index cases are associated to infection among contacts.

## Methods

### Study design and setting

A community-based cross-sectional study was conducted in the state of São Paulo, Brazil, from January 2010 to December 2020, using routinely collected data from the State Tuberculosis Control Program. São Paulo state is located in the Southeast region of Brazil and is the most populous and developed in the country. It leads the nation in TB cases, accounting for 24.5% of the total, with an estimated incidence above the national average of 42 cases per 100,000 person-years [12].

### Participants

The study included all contacts of new TB cases (index cases), defined as individuals who had never received TB treatment or had taken anti-TB medications for one month or less [1,11]. In this analysis, index cases were defined as patients diagnosed with TB according to national guidelines [11], and contacts were any individuals who had been exposed to an index case [7]. Screened contacts were those identified by index cases in the notification form. Examined contacts included all individuals who underwent clinical evaluation. During this assessment, it was expected that all steps proposed by the contact investigation algorithm would be completed.

### Data source

All information was obtained through the electronic Notification and Monitoring System for Tuberculosis Cases in the State of São Paulo (TBWEB). This system encompasses all TB cases reported by state residents and, in addition to the individual and clinical characteristics of index cases, includes three specific fields related to contact investigation, specifying the number of contacts screened, examined, and diagnosed with active TB per index patient.

### Contact investigation procedure

Following the diagnosis of TB in the index case, regardless of clinical presentation, healthcare professionals conduct an in-person interview with the patient to gather information about all their contacts, including names, ages, and risk assessment, in order to prioritize clinical examination. Furthermore, they educate the patient on the importance of contact investigation. Subsequently, they request that contacts visit the designated health facility for evaluation or contact the contacts to schedule a visit, as needed [7,11].

Contacts are then assessed for the presence of persistent cough of any duration or other symptoms such as persistent fever, weight loss, anorexia, and night sweats, among others. Regardless of symptoms presented, a chest X-ray is requested. Contacts under 10 years old undergo tuberculin skin testing or interferon-gamma release assay (IFN-γ) to check for LTBI. Those over 10 years old who are capable of producing a sputum sample are investigated using sputum smear microscopy or GeneXpert MTB/RIF®. Cases positive on these tests are diagnosed with active TB and immediately start treatment, tailored to drug resistance patterns [11].

Contacts unable to produce sputum or those with negative sputum results but abnormal radiographic findings are referred for additional medical clinical evaluation. Also, asymptomatic contacts are screened for LTBI and, if necessary, referred for treatment [11]. However, we did not have access to this information; therefore, our assessment was limited to cases of active TB. It is important to note that all tests for the diagnosis and treatment of TB are fully covered by the Brazilian Unified Health System (SUS) [11]. Supplementary materials provide flowcharts for contact investigation based on the age of contacts (Supplementary Figure S1 and S2).

### Variables

The primary outcome was the detection of TB among contacts of TB index patients. We interpreted "positive yield" as the prevalence of this outcome, i.e., the percentage of screened contacts diagnosed with active TB as a result of TB contact investigation strategy [7,13]. The following indicators were also assessed: proportion of index cases for which contacts were registered; proportion of screened contacts who were examined; number needed to screen (NNR), and number needed to test (NNT), to identify one new TB case [7].

Due to the absence of individual information on contacts, the independent variables considered in the analyses pertained to characteristics of the index cases. These included sociodemographic information, health behaviours, medical history, and TB-related characteristics. A full description of all variables used in the study can be found in the Supplementary Table S1.

### Statistical analyses

The characteristics of index cases and information about TB contact investigation were presented descriptively.

### Predictive model for contact examination

We conducted various predictive modelling to estimate the likelihood of index cases having their contacts examined based on their characteristics. The final model was selected based on multiple criteria (Supplementary Figure S4, and Table S2). The zero-inflated Poisson (ZIP) regression model showed the best fit compared to the other models evaluated and was used to obtain probability estimates (Figure 1). For more details, please refer to the supplementary material.

**Figure 1.**
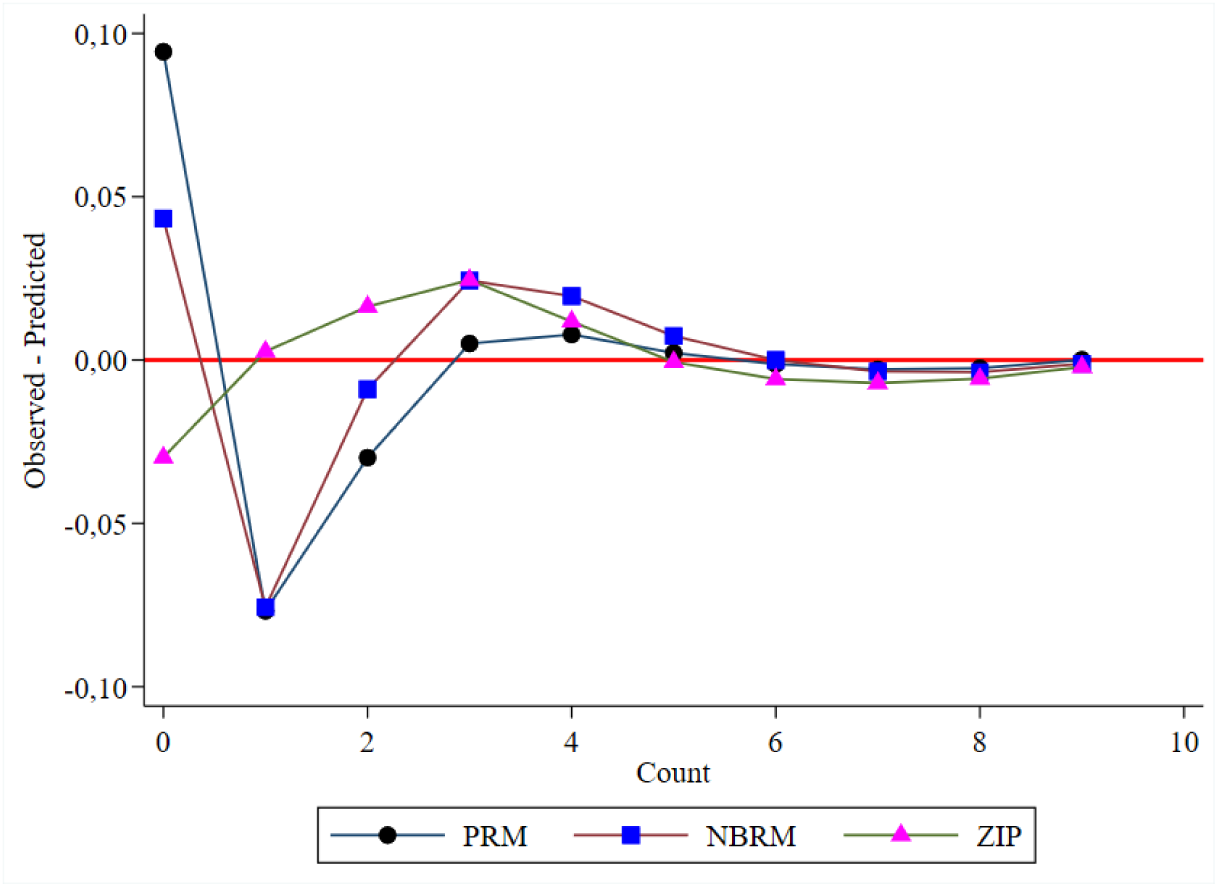
Comparisons among observed versus predicted probabilities among count models. Abbreviations: PRM – Poisson Regression Model. NBRM – Negative Binomial Regression Model, ZIP – Zero-inflated Poisson.

### Prevalence and factors associated with infection among contacts

We calculated sampling weights as the inverse probability of contacts being examined using the results from the previous predictive model. This approach allowed us to derive two prevalence estimates: one unweighted, representing the examined contacts, and one weighted, representing the screened contacts, both with their respective 95% confidence intervals (CI). By comparing these two values, we estimated underreporting, indicating the likely number of undetected cases among all screened contacts. It is important to note that there is no record explaining why not all screened contacts were examined. Therefore, we assumed that unexamined contacts do not have a lower prevalence of TB, considering the known characteristics of the index cases.

We investigated factors associated with TB among contacts based on index case characteristics using multilevel mixed-effects Poisson regression model. Additionally, we incorporated random effects at the municipal level to address variability not explained by fixed predictors and employed robust standard error estimates. We obtained adjusted models both unweighted and weighted, allowing estimates for examined contacts and the total screened contacts, respectively. Moreover, the use of weighting enabled us to avoid introducing collider bias into the analysis (Supplementary Figure S5). To address the missing values in the age variable, we performed simple data imputation (0.1% of index cases). For variables with more than 5% missing values, we included these cases as an additional category in the analysis.

The adjusted models were built using a hierarchical analysis, structured based on a conceptual framework (Supplementary Figure S6). This framework includes: first, temporal and geographical characteristics of index cases, as distal variables; second, sociodemographic and health characteristics, as intermediate I and II variables; and third, case detection strategies and clinical characteristics of index patients, as proximal variables. We interpreted the results in terms of prevalence ratio (PR) with their 95% CI, adopting a significance level of 5%.

We also estimated the intraclass correlation coefficient (ICC) for each multilevel Poisson regression model to assess the proportion of total variation in TB prevalence among contacts attributable to differences between municipalities. Furthermore, we examined the effect of each municipality on TB prevalence among contacts and generated a caterpillar plot that organizes predicted proportions in ascending order along with their respective 95% CIs.

All analyses were performed using Stata version 16.1 (StataCorp LP, College Station, Texas, USA). This study was reported according to the recommendations of the RECORD statement.

## Results

### Characteristics of index cases

Between January 1, 2010 and December 31, 2020, a total of 224,762 TB cases were reported to the TBWEB system. Of these index cases, 70.7% were male (n=131,777) with a median age of 35 years (IQR: 25-49), and more than half were from the metropolitan area of São Paulo (52.0%, n=96,975). Outpatient clinics passive case-finding (47.8%, n=89,111) was the most common detection strategy. In terms of diagnosis, the majority of cases showed abnormalities on chest X-ray (68.5%, n=127,729), pulmonary anatomical classification (84.6%, n=157,830), and positive bacteriological confirmation (68.1%, n=127,015). Table 1 shows the remaining characteristics of the index cases and compares them with the screening of at least one contact.

**Table 1.**
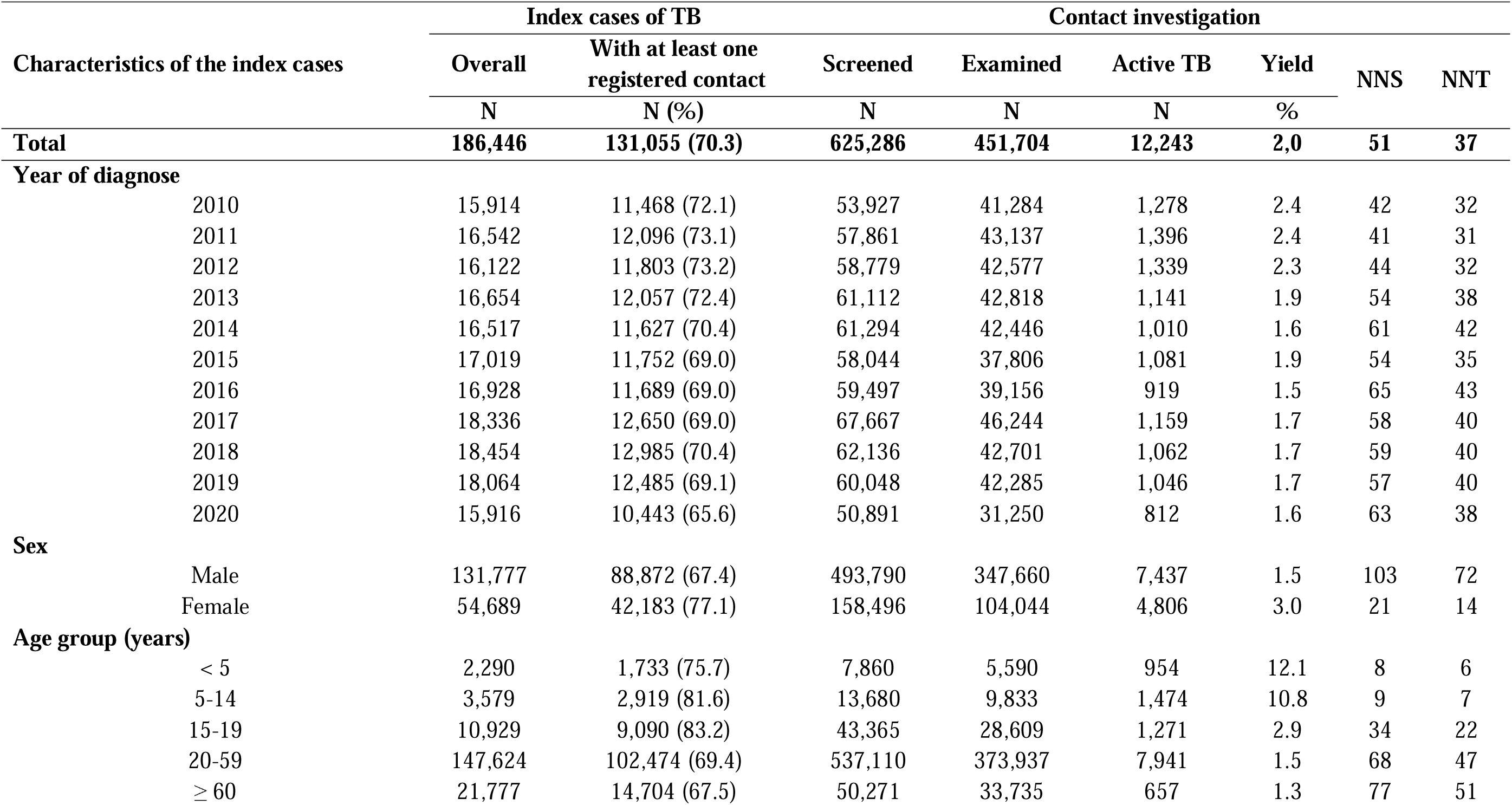
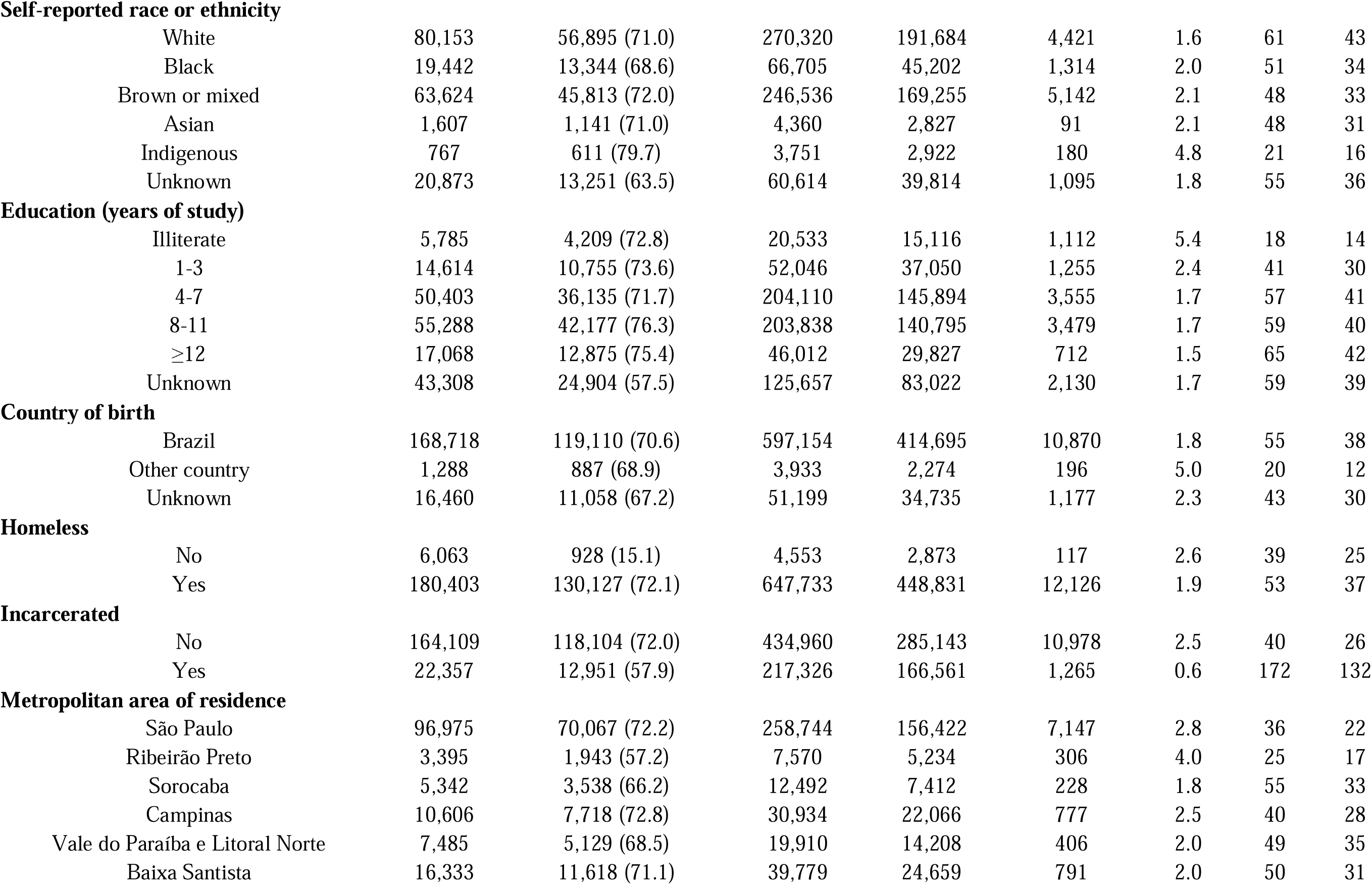
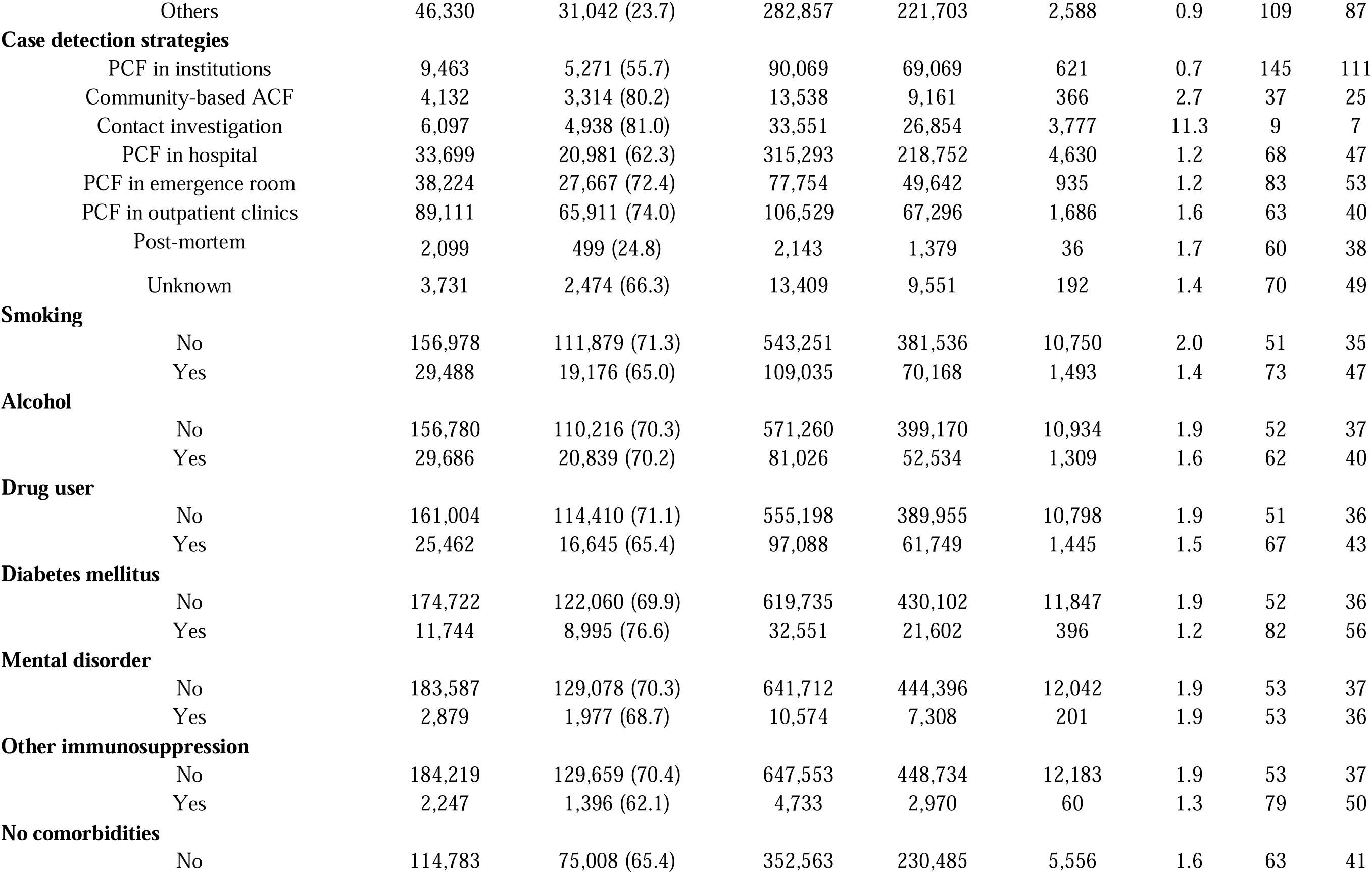
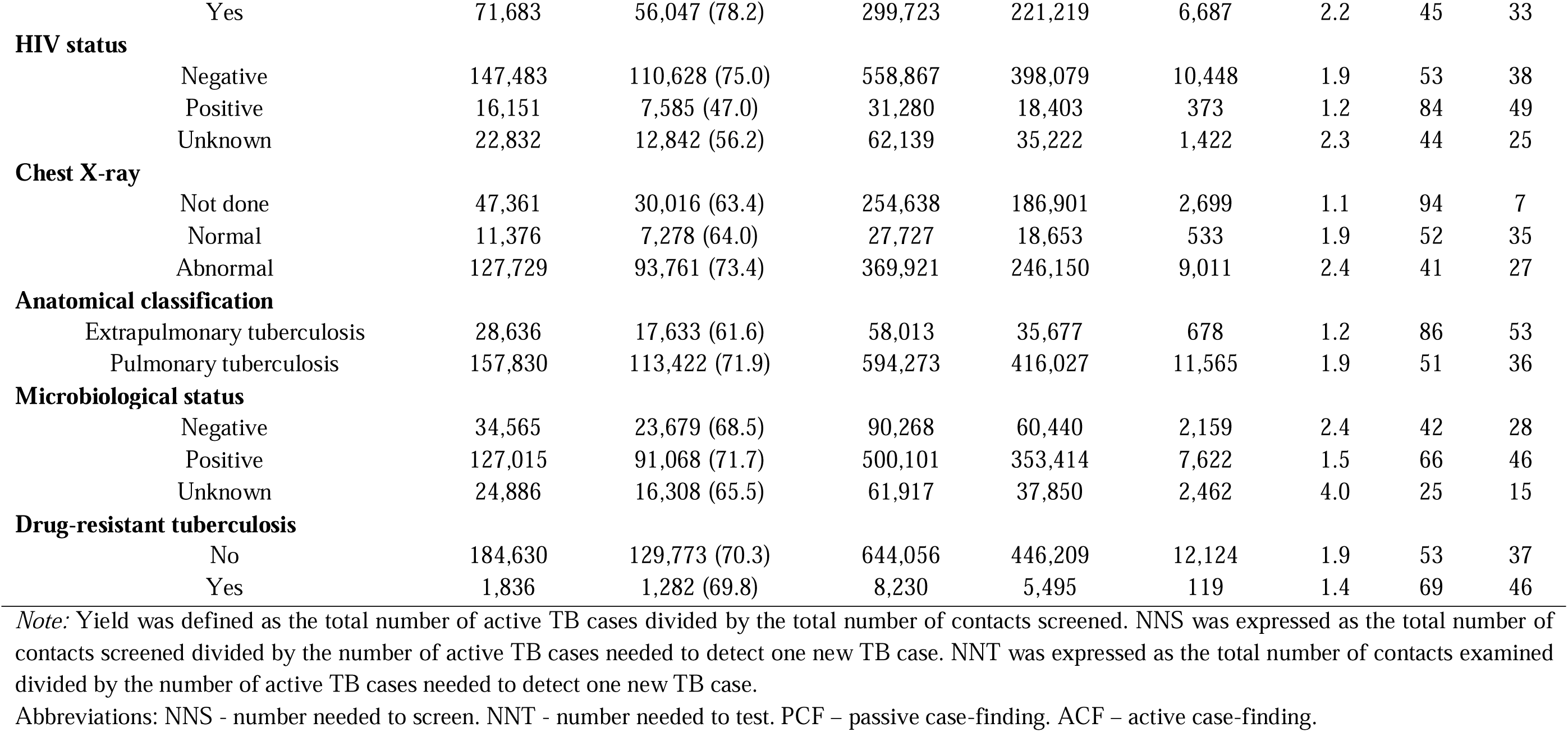
Characteristics of tuberculosis index cases, yield of tuberculosis contact investigations, number needed to screen, and number needed to treat in São Paulo, Brazil, 2010-2020.

| Characteristics of the index cases |  | Index cases of TB |  | Contact investigation |  |  |  |  |  |
| --- | --- | --- | --- | --- | --- | --- | --- | --- | --- |
|  |  | Overall | With at least one registered contact | Screened | Examined | Active TB | Yield | NNS | NNT |
| Total |  | 186,446 | 131,055 (70.3) | 625,286 | 451,704 | 12,243 | 2,0 | 51 | 37 |
| Year of diagnose |  |  |  |  |  |  |  |  |  |
|  | 2010 | 15,914 | 11,468 (72.1) | 53,927 | 41,284 | 1,278 | 2.4 | 42 | 32 |
|  | 2011 | 16,542 | 12,096 (73.1) | 57,861 | 43,137 | 1,396 | 2.4 | 41 | 31 |
|  | 2012 | 16,122 | 11,803 (73.2) | 58,779 | 42,577 | 1,339 | 2.3 | 44 | 32 |
|  | 2013 | 16,654 | 12,057 (72.4) | 61,112 | 42,818 | 1,141 | 1.9 | 54 | 38 |
|  | 2014 | 16,517 | 11,627 (70.4) | 61,294 | 42,446 | 1,010 | 1.6 | 61 | 42 |
|  | 2015 | 17,019 | 11,752 (69.0) | 58,044 | 37,806 | 1,081 | 1.9 | 54 | 35 |
|  | 2016 | 16,928 | 11,689 (69.0) | 59,497 | 39,156 | 919 | 1.5 | 65 | 43 |
|  | 2017 | 18,336 | 12,650 (69.0) | 67,667 | 46,244 | 1,159 | 1.7 | 58 | 40 |
|  | 2018 | 18,454 | 12,985 (70.4) | 62,136 | 42,701 | 1,062 | 1.7 | 59 | 40 |
|  | 2019 | 18,064 | 12,485 (69.1) | 60,048 | 42,285 | 1,046 | 1.7 | 57 | 40 |
|  | 2020 | 15,916 | 10,443 (65.6) | 50,891 | 31,250 | 812 | 1.6 | 63 | 38 |
| Sex |  |  |  |  |  |  |  |  |  |
|  | Male | 131,777 | 88,872 (67.4) | 493,790 | 347,660 | 7,437 | 1.5 | 103 | 72 |
|  | Female | 54,689 | 42,183 (77.1) | 158,496 | 104,044 | 4,806 | 3.0 | 21 | 14 |
| Age group (years) |  |  |  |  |  |  |  |  |  |
|  | < 5 | 2,290 | 1,733 (75.7) | 7,860 | 5,590 | 954 | 12.1 | 8 | 6 |
|  | 5-14 | 3,579 | 2,919 (81.6) | 13,680 | 9,833 | 1,474 | 10.8 | 9 | 7 |
|  | 15-19 | 10,929 | 9,090 (83.2) | 43,365 | 28,609 | 1,271 | 2.9 | 34 | 22 |
|  | 20-59 | 147,624 | 102,474 (69.4) | 537,110 | 373,937 | 7,941 | 1.5 | 68 | 47 |
|  | ≥ 60 | 21,777 | 14,704 (67.5) | 50,271 | 33,735 | 657 | 1.3 | 77 | 51 |

**Self-reported race or ethnicity**
|  |  |  |  |  |  |  |  |  |
| --- | --- | --- | --- | --- | --- | --- | --- | --- |
| White | 80,153 | 56,895 (71.0) | 270,320 | 191,684 | 4,421 | 1.6 | 61 | 43 |
| Black | 19,442 | 13,344 (68.6) | 66,705 | 45,202 | 1,314 | 2.0 | 51 | 34 |
| Brown or mixed | 63,624 | 45,813 (72.0) | 246,536 | 169,255 | 5,142 | 2.1 | 48 | 33 |
| Asian | 1,607 | 1,141 (71.0) | 4,360 | 2,827 | 91 | 2.1 | 48 | 31 |
| Indigenous | 767 | 611 (79.7) | 3,751 | 2,922 | 180 | 4.8 | 21 | 16 |
| Unknown | 20,873 | 13,251 (63.5) | 60,614 | 39,814 | 1,095 | 1.8 | 55 | 36 |

**Education (years of study)**
|  |  |  |  |  |  |  |  |  |
| --- | --- | --- | --- | --- | --- | --- | --- | --- |
| Illiterate | 5,785 | 4,209 (72.8) | 20,533 | 15,116 | 1,112 | 5.4 | 18 | 14 |
| 1-3 | 14,614 | 10,755 (73.6) | 52,046 | 37,050 | 1,255 | 2.4 | 41 | 30 |
| 4-7 | 50,403 | 36,135 (71.7) | 204,110 | 145,894 | 3,555 | 1.7 | 57 | 41 |
| 8-11 | 55,288 | 42,177 (76.3) | 203,838 | 140,795 | 3,479 | 1.7 | 59 | 40 |
| ≥12 | 17,068 | 12,875 (75.4) | 46,012 | 29,827 | 712 | 1.5 | 65 | 42 |
| Unknown | 43,308 | 24,904 (57.5) | 125,657 | 83,022 | 2,130 | 1.7 | 59 | 39 |

**Country of birth**
|  |  |  |  |  |  |  |  |  |
| --- | --- | --- | --- | --- | --- | --- | --- | --- |
| Brazil | 168,718 | 119,110 (70.6) | 597,154 | 414,695 | 10,870 | 1.8 | 55 | 38 |
| Other country | 1,288 | 887 (68.9) | 3,933 | 2,274 | 196 | 5.0 | 20 | 12 |
| Unknown | 16,460 | 11,058 (67.2) | 51,199 | 34,735 | 1,177 | 2.3 | 43 | 30 |

**Homeless**
|  |  |  |  |  |  |  |  |  |
| --- | --- | --- | --- | --- | --- | --- | --- | --- |
| No | 6,063 | 928 (15.1) | 4,553 | 2,873 | 117 | 2.6 | 39 | 25 |
| Yes | 180,403 | 130,127 (72.1) | 647,733 | 448,831 | 12,126 | 1.9 | 53 | 37 |

**Incarcerated**
|  |  |  |  |  |  |  |  |  |
| --- | --- | --- | --- | --- | --- | --- | --- | --- |
| No | 164,109 | 118,104 (72.0) | 434,960 | 285,143 | 10,978 | 2.5 | 40 | 26 |
| Yes | 22,357 | 12,951 (57.9) | 217,326 | 166,561 | 1,265 | 0.6 | 172 | 132 |

**Metropolitan area of residence**
|  |  |  |  |  |  |  |  |  |
| --- | --- | --- | --- | --- | --- | --- | --- | --- |
| São Paulo | 96,975 | 70,067 (72.2) | 258,744 | 156,422 | 7,147 | 2.8 | 36 | 22 |
| Ribeirão Preto | 3,395 | 1,943 (57.2) | 7,570 | 5,234 | 306 | 4.0 | 25 | 17 |
| Sorocaba | 5,342 | 3,538 (66.2) | 12,492 | 7,412 | 228 | 1.8 | 55 | 33 |
| Campinas | 10,606 | 7,718 (72.8) | 30,934 | 22,066 | 777 | 2.5 | 40 | 28 |
| Vale do Paraíba e Litoral Norte | 7,485 | 5,129 (68.5) | 19,910 | 14,208 | 406 | 2.0 | 49 | 35 |
| Baixa Santista | 16,333 | 11,618 (71.1) | 39,779 | 24,659 | 791 | 2.0 | 50 | 31 |
| Others | 46,330 | 31,042 (23.7) | 282,857 | 221,703 | 2,588 | 0.9 | 109 | 87 |
| **Case detection strategies** |  |  |  |  |  |  |  |  |
| PCF in institutions | 9,463 | 5,271 (55.7) | 90,069 | 69,069 | 621 | 0.7 | 145 | 111 |
| Community-based ACF | 4,132 | 3,314 (80.2) | 13,538 | 9,161 | 366 | 2.7 | 37 | 25 |
| Contact investigation | 6,097 | 4,938 (81.0) | 33,551 | 26,854 | 3,777 | 11.3 | 9 | 7 |
| PCF in hospital | 33,699 | 20,981 (62.3) | 315,293 | 218,752 | 4,630 | 1.2 | 68 | 47 |
| PCF in emergence room | 38,224 | 27,667 (72.4) | 77,754 | 49,642 | 935 | 1.2 | 83 | 53 |
| PCF in outpatient clinics | 89,111 | 65,911 (74.0) | 106,529 | 67,296 | 1,686 | 1.6 | 63 | 40 |
| Post-mortem | 2,099 | 499 (24.8) | 2,143 | 1,379 | 36 | 1.7 | 60 | 38 |
| Unknown | 3,731 | 2,474 (66.3) | 13,409 | 9,551 | 192 | 1.4 | 70 | 49 |
| **Smoking** |  |  |  |  |  |  |  |  |
| No | 156,978 | 111,879 (71.3) | 543,251 | 381,536 | 10,750 | 2.0 | 51 | 35 |
| Yes | 29,488 | 19,176 (65.0) | 109,035 | 70,168 | 1,493 | 1.4 | 73 | 47 |
| **Alcohol** |  |  |  |  |  |  |  |  |
| No | 156,780 | 110,216 (70.3) | 571,260 | 399,170 | 10,934 | 1.9 | 52 | 37 |
| Yes | 29,686 | 20,839 (70.2) | 81,026 | 52,534 | 1,309 | 1.6 | 62 | 40 |
| **Drug user** |  |  |  |  |  |  |  |  |
| No | 161,004 | 114,410 (71.1) | 555,198 | 389,955 | 10,798 | 1.9 | 51 | 36 |
| Yes | 25,462 | 16,645 (65.4) | 97,088 | 61,749 | 1,445 | 1.5 | 67 | 43 |
| **Diabetes mellitus** |  |  |  |  |  |  |  |  |
| No | 174,722 | 122,060 (69.9) | 619,735 | 430,102 | 11,847 | 1.9 | 52 | 36 |
| Yes | 11,744 | 8,995 (76.6) | 32,551 | 21,602 | 396 | 1.2 | 82 | 56 |
| **Mental disorder** |  |  |  |  |  |  |  |  |
| No | 183,587 | 129,078 (70.3) | 641,712 | 444,396 | 12,042 | 1.9 | 53 | 37 |
| Yes | 2,879 | 1,977 (68.7) | 10,574 | 7,308 | 201 | 1.9 | 53 | 36 |
| **Other immunosuppression** |  |  |  |  |  |  |  |  |
| No | 184,219 | 129,659 (70.4) | 647,553 | 448,734 | 12,183 | 1.9 | 53 | 37 |
| Yes | 2,247 | 1,396 (62.1) | 4,733 | 2,970 | 60 | 1.3 | 79 | 50 |
| **No comorbidities** |  |  |  |  |  |  |  |  |
| No | 114,783 | 75,008 (65.4) | 352,563 | 230,485 | 5,556 | 1.6 | 63 | 41 |
|  | Yes | 71,683 | 56,047 (78.2) | 299,723 | 221,219 | 6,687 | 2.2 | 45 | 33 |
| **HIV status** |  |  |  |  |  |  |  |  |  |
|  | Negative | 147,483 | 110,628 (75.0) | 558,867 | 398,079 | 10,448 | 1.9 | 53 | 38 |
|  | Positive | 16,151 | 7,585 (47.0) | 31,280 | 18,403 | 373 | 1.2 | 84 | 49 |
|  | Unknown | 22,832 | 12,842 (56.2) | 62,139 | 35,222 | 1,422 | 2.3 | 44 | 25 |
| **Chest X-ray** |  |  |  |  |  |  |  |  |  |
|  | Not done | 47,361 | 30,016 (63.4) | 254,638 | 186,901 | 2,699 | 1.1 | 94 | 7 |
|  | Normal | 11,376 | 7,278 (64.0) | 27,727 | 18,653 | 533 | 1.9 | 52 | 35 |
|  | Abnormal | 127,729 | 93,761 (73.4) | 369,921 | 246,150 | 9,011 | 2.4 | 41 | 27 |
| **Anatomical classification** |  |  |  |  |  |  |  |  |  |
|  | Extrapulmonary tuberculosis | 28,636 | 17,633 (61.6) | 58,013 | 35,677 | 678 | 1.2 | 86 | 53 |
|  | Pulmonary tuberculosis | 157,830 | 113,422 (71.9) | 594,273 | 416,027 | 11,565 | 1.9 | 51 | 36 |
| **Microbiological status** |  |  |  |  |  |  |  |  |  |
|  | Negative | 34,565 | 23,679 (68.5) | 90,268 | 60,440 | 2,159 | 2.4 | 42 | 28 |
|  | Positive | 127,015 | 91,068 (71.7) | 500,101 | 353,414 | 7,622 | 1.5 | 66 | 46 |
|  | Unknown | 24,886 | 16,308 (65.5) | 61,917 | 37,850 | 2,462 | 4.0 | 25 | 15 |
| **Drug-resistant tuberculosis** |  |  |  |  |  |  |  |  |  |
|  | No | 184,630 | 129,773 (70.3) | 644,056 | 446,209 | 12,124 | 1.9 | 53 | 37 |
|  | Yes | 1,836 | 1,282 (69.8) | 8,230 | 5,495 | 119 | 1.4 | 69 | 46 |

*Note:* Yield was defined as the total number of active TB cases divided by the total number of contacts screened. NNS was expressed as the total number of contacts screened divided by the number of active TB cases needed to detect one new TB case. NNT was expressed as the total number of contacts examined divided by the number of active TB cases needed to detect one new TB case.
Abbreviations: NNS - number needed to screen. NNT - number needed to test. PCF – passive case-finding. ACF – active case-finding.

### The yield of contact investigation

Among index cases, 131,055 (70.3%) had at least one contact registered in TBWEB, totalling 652,286 contacts screened (5 per index case). The median number of contacts screened per index case was 3 (IQR: 2-5), ranging from 1 to 300. Regarding contact investigation, 451,704 (69.2%) underwent examinations to detect the presence of the disease. The median number of contacts examined per index case was 3 (IQR: 2-5), ranging from 1 to 272. In total, 12,243 new TB cases were diagnosed, representing an overall yield of contact investigation of approximately 2.0% (Figure 2), resulting in an NNS of 51 and NNT of 37 (Table 1). This yield was higher during the period from 2010 to 2012, reaching a proportion of 2.4% (NNS=42), and varied from 0.6% (NNS=172) among contacts of index cases in correctional facilities to 12.1% (NNS=8) among contacts of index cases under 5 years old. Table 1 shows the yield, NNR, and NNT values according to other characteristics of index cases.

**Figure 2.**
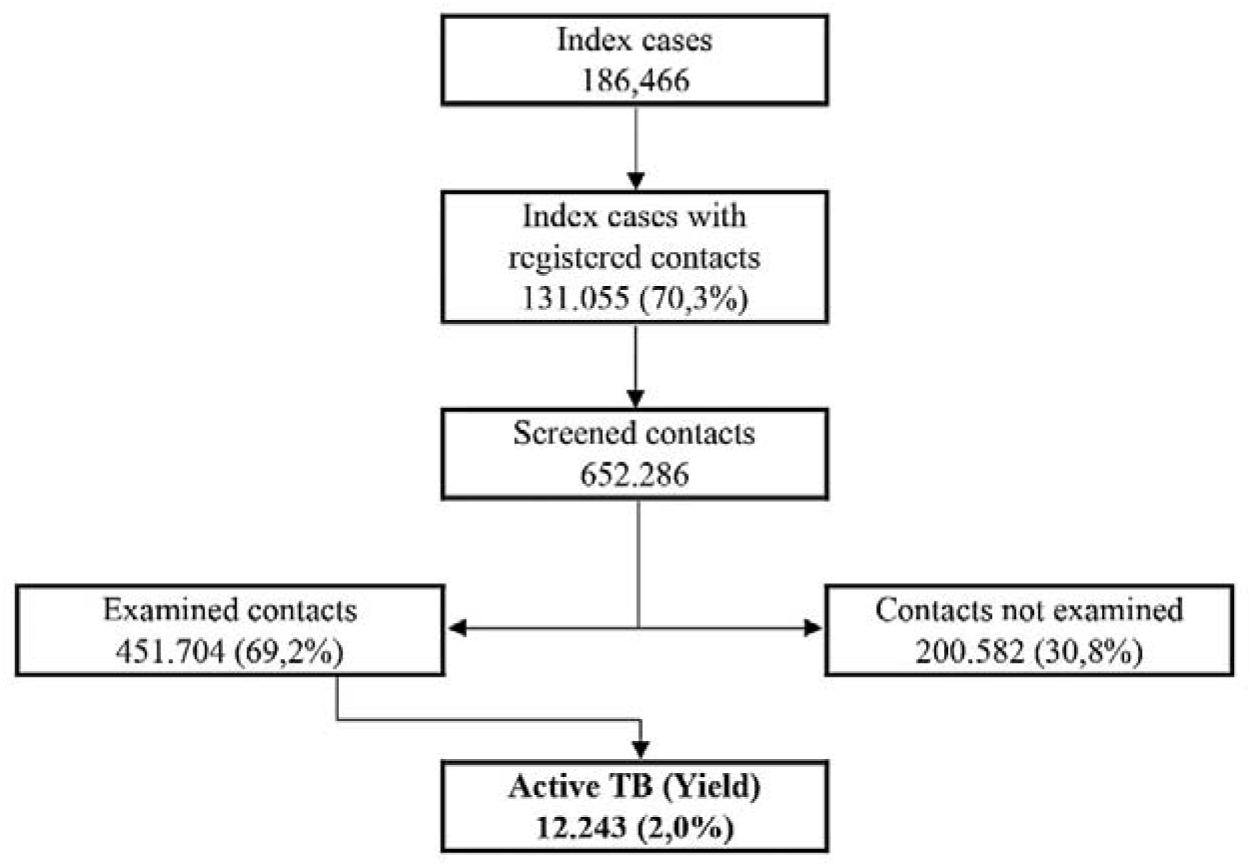
Caterpillar plot showing the effect of municipalities on tuberculosis prevalence among contacts and their respective 95% confidence intervals (n=639). São Paulo, Brazil, 2010-2020.

### The prevalence of TB among contacts

We found that the overall proportion of TB among examined contacts was 2.7% (95% CI: 2.6%–2.8%). The weighted prevalence for the total screened contacts was 2.8% (95% CI: 2.7%–2.9%), resulting in 18,264 cases (95% CI: 17,612–18,916) among all screened contacts. These results suggest a possible underreporting of 6,021 cases (95% CI: 5,269–6,673) of undetected infections among contacts referred by index cases. Table 2 shows additional prevalence values disaggregated according to characteristics of the index cases.

**Table 2.**
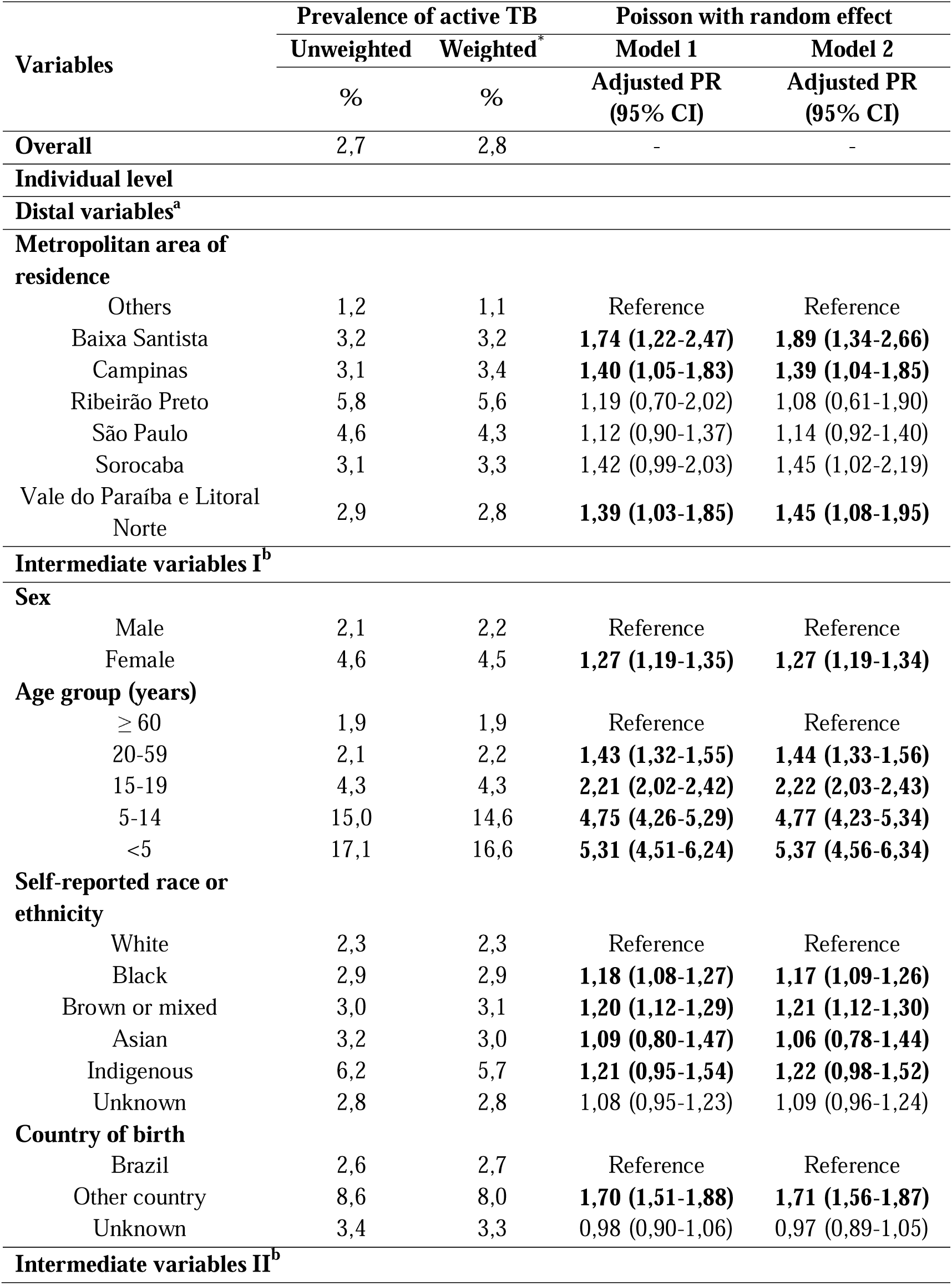
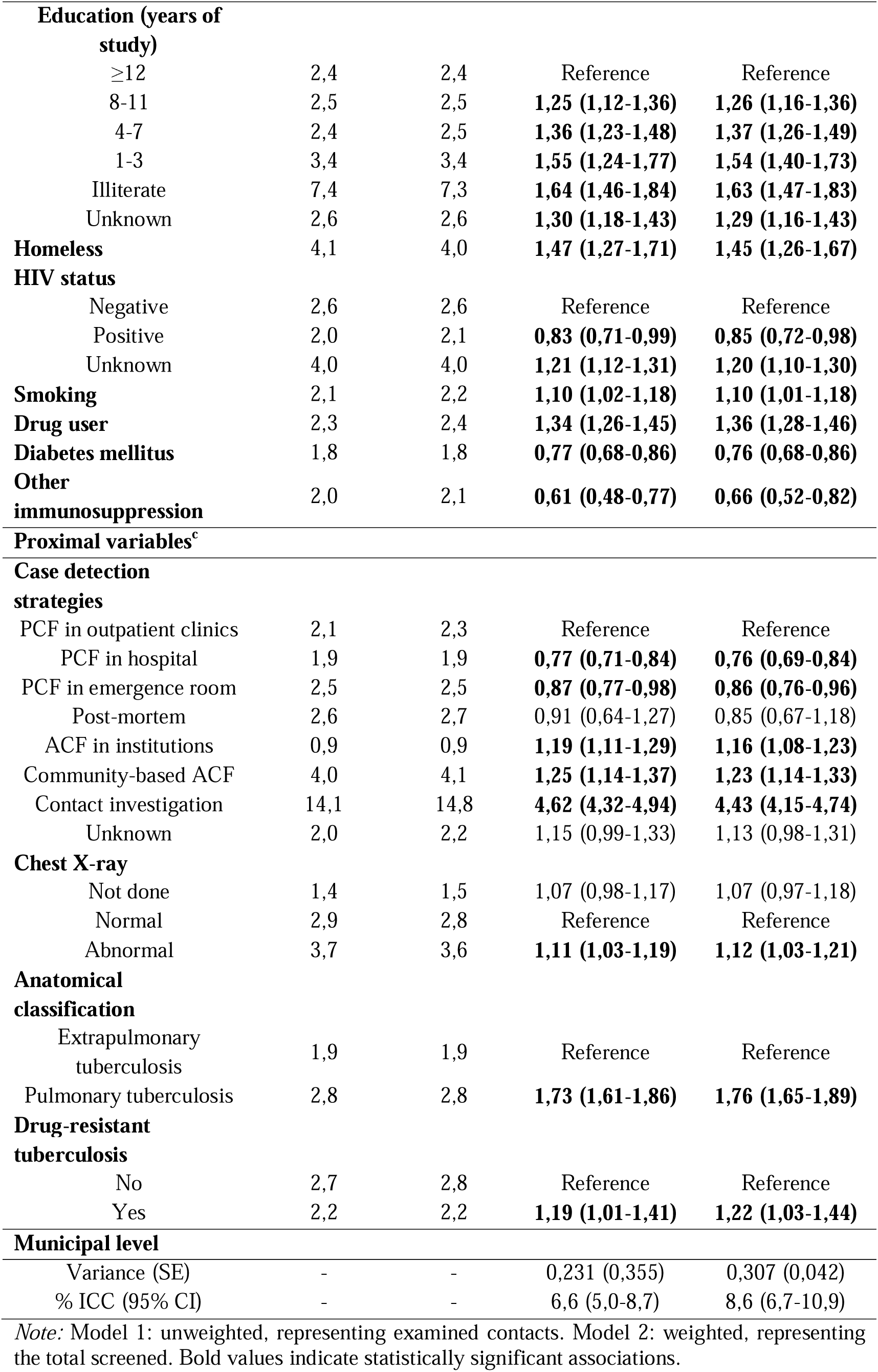
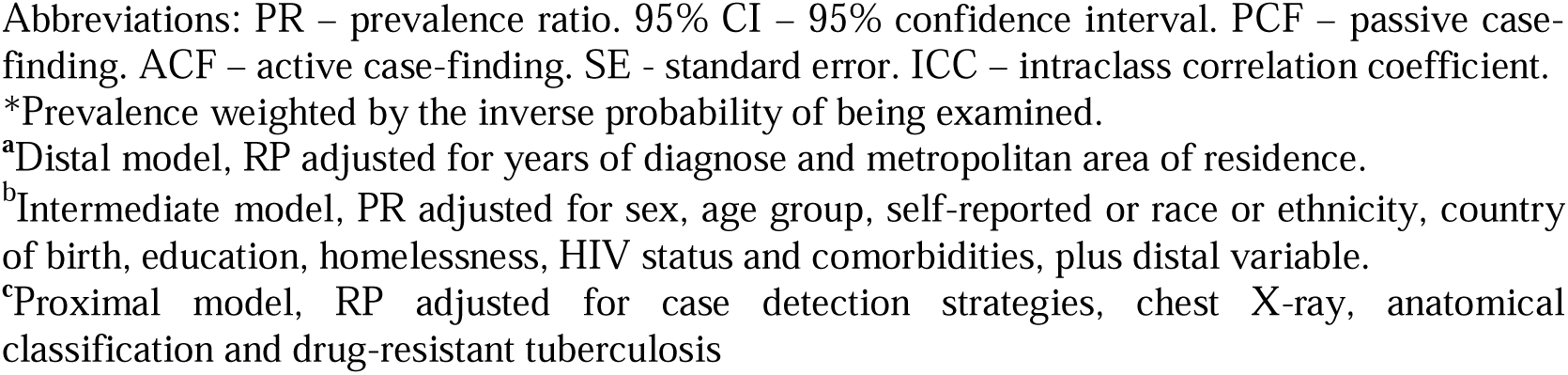
Prevalences and multilevel Poisson regression analysis adjusted for characteristics of tuberculosis index cases associated with the presence of tuberculosis diagnosis among their contacts. São Paulo, Brazil, 2010-2020.

| Variables | Prevalence of active TB |  | Poisson with random effect |  |
| --- | --- | --- | --- | --- |
|  | Unweighted | Weighted* | Model 1 | Model 2 |
|  | % | % | Adjusted PR<br>(95% CI) | Adjusted PR<br>(95% CI) |
| **Overall** | 2,7 | 2,8 | - | - |
| **Individual level** |  |  |  |  |
| **Distal variables<sup>a</sup>** |  |  |  |  |
| **Metropolitan area of residence** |  |  |  |  |
| Others | 1,2 | 1,1 | Reference | Reference |
| Baixa Santista | 3,2 | 3,2 | **1,74 (1,22-2,47)** | **1,89 (1,34-2,66)** |
| Campinas | 3,1 | 3,4 | **1,40 (1,05-1,83)** | **1,39 (1,04-1,85)** |
| Ribeirão Preto | 5,8 | 5,6 | 1,19 (0,70-2,02) | 1,08 (0,61-1,90) |
| São Paulo | 4,6 | 4,3 | 1,12 (0,90-1,37) | 1,14 (0,92-1,40) |
| Sorocaba | 3,1 | 3,3 | 1,42 (0,99-2,03) | 1,45 (1,02-2,19) |
| Vale do Paraíba e Litoral Norte | 2,9 | 2,8 | **1,39 (1,03-1,85)** | **1,45 (1,08-1,95)** |
| **Intermediate variables I<sup>b</sup>** |  |  |  |  |
| **Sex** |  |  |  |  |
| Male | 2,1 | 2,2 | Reference | Reference |
| Female | 4,6 | 4,5 | **1,27 (1,19-1,35)** | **1,27 (1,19-1,34)** |
| **Age group (years)** |  |  |  |  |
| ≥ 60 | 1,9 | 1,9 | Reference | Reference |
| 20-59 | 2,1 | 2,2 | **1,43 (1,32-1,55)** | **1,44 (1,33-1,56)** |
| 15-19 | 4,3 | 4,3 | **2,21 (2,02-2,42)** | **2,22 (2,03-2,43)** |
| 5-14 | 15,0 | 14,6 | **4,75 (4,26-5,29)** | **4,77 (4,23-5,34)** |
| <5 | 17,1 | 16,6 | **5,31 (4,51-6,24)** | **5,37 (4,56-6,34)** |
| **Self-reported race or ethnicity** |  |  |  |  |
| White | 2,3 | 2,3 | Reference | Reference |
| Black | 2,9 | 2,9 | **1,18 (1,08-1,27)** | **1,17 (1,09-1,26)** |
| Brown or mixed | 3,0 | 3,1 | **1,20 (1,12-1,29)** | **1,21 (1,12-1,30)** |
| Asian | 3,2 | 3,0 | **1,09 (0,80-1,47)** | **1,06 (0,78-1,44)** |
| Indigenous | 6,2 | 5,7 | **1,21 (0,95-1,54)** | **1,22 (0,98-1,52)** |
| Unknown | 2,8 | 2,8 | 1,08 (0,95-1,23) | 1,09 (0,96-1,24) |
| **Country of birth** |  |  |  |  |
| Brazil | 2,6 | 2,7 | Reference | Reference |
| Other country | 8,6 | 8,0 | **1,70 (1,51-1,88)** | **1,71 (1,56-1,87)** |
| Unknown | 3,4 | 3,3 | 0,98 (0,90-1,06) | 0,97 (0,89-1,05) |
| **Intermediate variables II<sup>b</sup>** |  |  |  |  |
| **Education (years of study)** |  |  |  |  |
| ≥12 | 2,4 | 2,4 | Reference | Reference |
| 8-11 | 2,5 | 2,5 | **1,25 (1,12-1,36)** | **1,26 (1,16-1,36)** |
| 4-7 | 2,4 | 2,5 | **1,36 (1,23-1,48)** | **1,37 (1,26-1,49)** |
| 1-3 | 3,4 | 3,4 | **1,55 (1,24-1,77)** | **1,54 (1,40-1,73)** |
| Illiterate | 7,4 | 7,3 | **1,64 (1,46-1,84)** | **1,63 (1,47-1,83)** |
| Unknown | 2,6 | 2,6 | **1,30 (1,18-1,43)** | **1,29 (1,16-1,43)** |
| **Homeless** | 4,1 | 4,0 | **1,47 (1,27-1,71)** | **1,45 (1,26-1,67)** |
| **HIV status** |  |  |  |  |
| Negative | 2,6 | 2,6 | Reference | Reference |
| Positive | 2,0 | 2,1 | **0,83 (0,71-0,99)** | **0,85 (0,72-0,98)** |
| Unknown | 4,0 | 4,0 | **1,21 (1,12-1,31)** | **1,20 (1,10-1,30)** |
| **Smoking** | 2,1 | 2,2 | **1,10 (1,02-1,18)** | **1,10 (1,01-1,18)** |
| **Drug user** | 2,3 | 2,4 | **1,34 (1,26-1,45)** | **1,36 (1,28-1,46)** |
| **Diabetes mellitus** | 1,8 | 1,8 | **0,77 (0,68-0,86)** | **0,76 (0,68-0,86)** |
| **Other immunosuppression** | 2,0 | 2,1 | **0,61 (0,48-0,77)** | **0,66 (0,52-0,82)** |
| **Proximal variables<sup>c</sup>** |  |  |  |  |
| **Case detection strategies** |  |  |  |  |
| PCF in outpatient clinics | 2,1 | 2,3 | Reference | Reference |
| PCF in hospital | 1,9 | 1,9 | **0,77 (0,71-0,84)** | **0,76 (0,69-0,84)** |
| PCF in emergence room | 2,5 | 2,5 | **0,87 (0,77-0,98)** | **0,86 (0,76-0,96)** |
| Post-mortem | 2,6 | 2,7 | 0,91 (0,64-1,27) | 0,85 (0,67-1,18) |
| ACF in institutions | 0,9 | 0,9 | **1,19 (1,11-1,29)** | **1,16 (1,08-1,23)** |
| Community-based ACF | 4,0 | 4,1 | **1,25 (1,14-1,37)** | **1,23 (1,14-1,33)** |
| Contact investigation | 14,1 | 14,8 | **4,62 (4,32-4,94)** | **4,43 (4,15-4,74)** |
| Unknown | 2,0 | 2,2 | 1,15 (0,99-1,33) | 1,13 (0,98-1,31) |
| **Chest X-ray** |  |  |  |  |
| Not done | 1,4 | 1,5 | 1,07 (0,98-1,17) | 1,07 (0,97-1,18) |
| Normal | 2,9 | 2,8 | Reference | Reference |
| Abnormal | 3,7 | 3,6 | **1,11 (1,03-1,19)** | **1,12 (1,03-1,21)** |
| **Anatomical classification** |  |  |  |  |
| Extrapulmonary tuberculosis | 1,9 | 1,9 | Reference | Reference |
| Pulmonary tuberculosis | 2,8 | 2,8 | **1,73 (1,61-1,86)** | **1,76 (1,65-1,89)** |
| **Drug-resistant tuberculosis** |  |  |  |  |
| No | 2,7 | 2,8 | Reference | Reference |
| Yes | 2,2 | 2,2 | **1,19 (1,01-1,41)** | **1,22 (1,03-1,44)** |
| **Municipal level** |  |  |  |  |
| Variance (SE) | - | - | 0,231 (0,355) | 0,307 (0,042) |
| % ICC (95% CI) | - | - | 6,6 (5,0-8,7) | 8,6 (6,7-10,9) |

*Note:* Model 1: unweighted, representing examined contacts. Model 2: weighted, representing the total screened. Bold values indicate statistically significant associations.
Abbreviations: PR – prevalence ratio, 95% CI – 95% confidence interval. PCF – passive case-finding, ACF – active case-finding, SE - standard error, ICC – intraclass correlation coefficient.
\*Prevalence weighted by the inverse probability of being examined.
<sup>a</sup>Distal model, RP adjusted for years of diagnose and metropolitan area of residence.
<sup>b</sup>Intermediate model, PR adjusted for sex, age group, self-reported or race or ethnicity, country of birth, education, homelessness, HIV status and comorbidities, plus distal variable.
<sup>c</sup>Proximal model, RP adjusted for case detection strategies, chest X-ray, anatomical classification and drug-resistant tuberculosis

### Factors associated with TB diagnosis among contacts

Index patients identified in the metropolitan areas of Campinas, Baixada Santista, Vale do Paraíba e Litoral Norte, and Sorocaba were associated with a higher likelihood of TB diagnosis among their contacts. This trend was also observed among contacts of female index patients, those of Black or Brown races/ethnicities, foreigners, and individuals experiencing homelessness. However, an inverse relationship was noted between the age and years of schooling of index patients and the likelihood of TB diagnosis among their contacts (Table 2).

Additionally, index cases with unknown HIV status, smokers, and illicit drug users also showed an increased probability of TB diagnosis among their contacts. Furthermore, index patients identified through active case-finding strategies, with abnormal chest X-ray results, pulmonary TB diagnosis, and drug resistance, were associated with a higher probability of TB diagnosis among their contacts (Table 2).

### Examining the effect of municipalities on TB prevalence among contacts

We found that 6.6% (95% CI: 5.0%–8.7%) and 8.6% (95% CI: 6.7%–10.9%) of the variation in TB prevalence among examined and screened contacts, respectively, was attributed to variation between municipalities (Table 2). The Figure 3 shows a caterpillar plot presenting estimated residuals for TB prevalence among contacts for the 639 municipalities along with their respective 95% CIs. For twenty-one municipalities, the 95% ICs were below the zero line, indicating lower predicted TB prevalence among contacts compared to the average. In contrast, forty-three municipalities had 95% CIs above the zero line, suggesting higher TB prevalence among contacts than the average. For 90% of the municipalities included, it was not possible to distinguish from the overall average due to overlapping 95% ICs with the zero line. For more information, please consult Supplementary Table S3.

**Figure 3.**
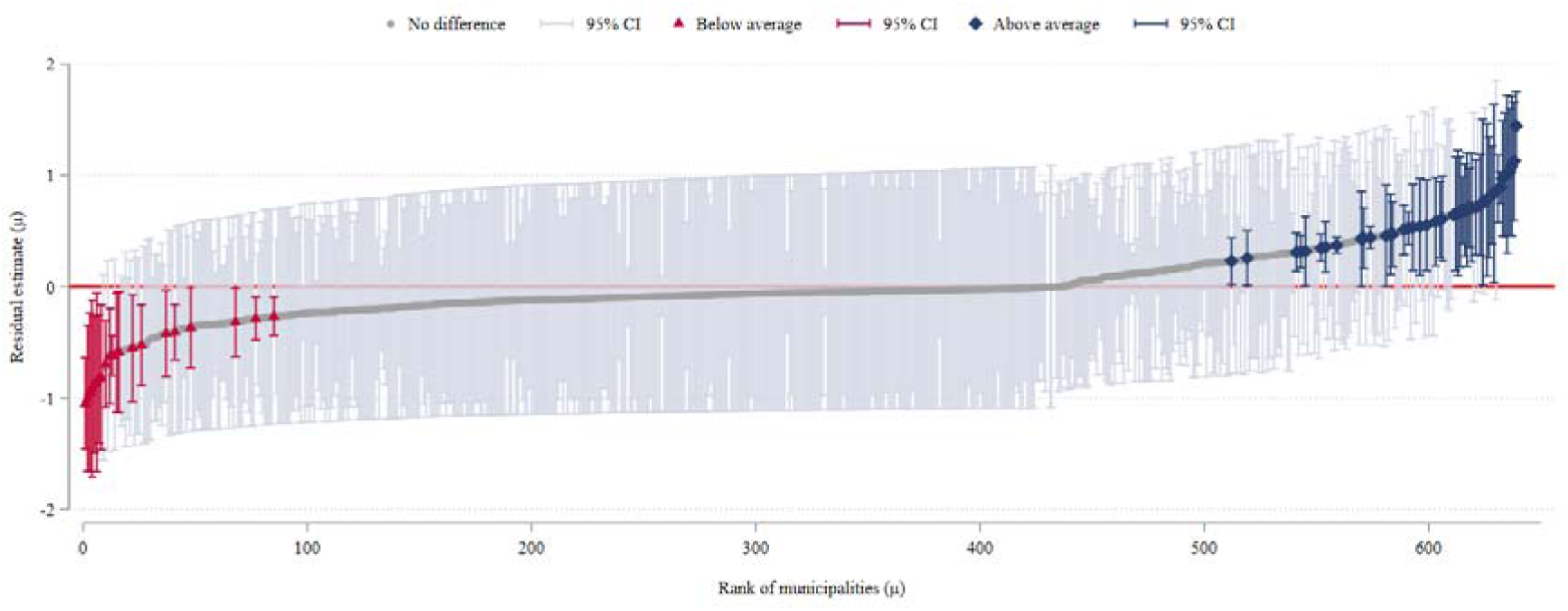
Caterpillar plot showing the effect of municipalities on tuberculosis prevalence among contacts and their respective 95% confidence intervals (n=639). São Paulo, Brazil, 2010-2020.

## Discussion

In this study, we evaluated the yield of TB contact investigation strategies among index cases in the state of São Paulo, Brazil, using available surveillance data. We estimated the prevalence of TB among contacts and identified the characteristics of index cases associated with active TB diagnosed among their contacts. The yield was approximately 2.0% among screened contacts, increasing to 2.7% among those examined. These results align with estimates from other studies in Brazil across different populations, ranging from 1.9% to 3.0% [14,15], although they remain slightly lower than the global range of 2.87% to 3.60% reported in previous studies [3,5,8,16]. Nevertheless, these numbers are comparable to those observed in other countries in the Americas (2.68%) [3] and in similar income and incidence settings (2.22% and 1.9%, respectively) [3,8]. They are also consistent with the yield reported in another study that used data from national TB surveillance program data (1.8%) [17]. Additionally, our findings reveal that the weighted prevalence considering all screened contacts was 2.8% (95% CI: 2,7%–2,9%), indicating a potential underreporting of 6,021 TB cases (95% CI: 5,269– 6,673), which represents for nearly one-third of all cases among contacts. This result supports with previous studies that used different approaches to determine underreporting [18].

Our study shows that, on average, 9 contacts are diagnosed with TB for every 100 index cases screened. This estimate likely underestimates the true proportion of cases per index patient, as 30% of them did not report any contacts. However, it is crucial to note that this non-screening rate is lower than that found in other high TB burden countries in Africa, Asia, and the Middle East [17]. Another significant finding is the proportion of contacts that were actually examined, which accounts for nearly 70% of the total contacts screened. We observed that this reduction in the number of contacts between the screening and testing stages may have contributed to case losses, which could partly explain the lower yield percentage found compared to other studies.

It is essential to emphasize that the possibility of underdiagnosis of the disease requires further investigation, especially due to previous findings that highlighted the stigma associated with TB and HIV as a significant barrier to contact investigation [19,20]. Furthermore, other studies indicates that difficulties in reporting contacts by index cases may be attributed not only to stigma but also to the complexity arising from the number and identity of potentially exposed contacts, the strategies used for contact tracing, limited knowledge about the disease among contacts, challenges in accessing health services, and inadequate follow-up by health teams after contact identification, among others [21,22]. These factors, whether alone or in combination, may contribute to a scenario where TB cases among contacts are not promptly diagnosed and treated, thereby increasing disease transmission [20].

This result further underscores the need to ensure the completion of the entire contact tracing cascade for all eligible index cases, thereby avoiding selection bias towards individuals who self-identify as symptomatic [23]. This process is crucial, particularly in resource-limited settings where contacts are encouraged to seek medical assistance only when symptoms appear [16]. Previous evidence supports this finding, demonstrating that locations testing all contacts, regardless of symptoms, achieved a more significant detection of TB cases compared to those applying more restrictive criteria [7,24]. Moreover, integrating laboratory tests into contact tracing activities can be a valuable investment, given the limitations of relying solely on symptoms to guide TB case screening [16,25]. It is also crucial to highlight the importance of identifying and initiating early treatment for LTBI among eligible contacts of index cases, which helps prevent future reactivations of TB [26]. Unfortunately, these data were not available for analysis in our study.

In this investigation, on average, 51 contacts need to be screened and 37 examined to identify a case of active TB, a result similar to other studies [17,27,28]. This implies conducting approximately ten home visits, considering the average number of contacts per index case (51/5), which can be a significant expenditure of resources and a burden on healthcare professionals, depending on the location. Notably, we observed that this number is significantly lower in certain groups, which supports World Health Organization (WHO) and national guidelines to target screening towards specific groups at higher risk of the disease [7,11]. This targeted approach can optimise resources and enhance TB control effectiveness, enabling earlier and more efficient disease detection, thereby contributing to reducing its burden and advancing towards its elimination [16,27].

Our analyses indicated that contacts of index cases with drug resistance were more likely to be diagnosed with TB compared to contacts of drug-susceptible index cases. While some studies, including our own, have observed this pattern of resistance [29], the literature still shows inconsistencies in the relationship between contacts and their index cases, highlighting the need for further studies [30]. We also observed that contacts of index cases with pulmonary TB and abnormalities on chest X-rays were more likely to be diagnosed with TB compared to cases with extrapulmonary TB and normal chest X-rays [9,14]. In addition, the strategy used to identify the TB index case was associated with the presence of the disease among contacts. This finding corroborates with others studies that emphasizes the effectiveness of active case-finding strategies in early detection of new cases and LTBI [5,31], and in reducing mortality and unfavourable treatment outcomes [6].

Studies have shown that smoking, illicit drug use, and other immunosuppressive conditions are known risk factors for TB infection, due to their negative impact on lung function and the immune system’s ability to fight infections [32,33]. In our investigation, we identified an association between TB index cases with such characteristics and the disease prevalence among their contacts [34,35]. We also noted an intriguing finding, contacts of TB index cases with diabetes mellitus (DM) showed a lower likelihood of TB diagnosis compared to those without DM, which concurs with previous studies [30]. However, this result contrasts with two other studies conducted in Brazil [34,36]. This discrepancy reveals the complexity of the relationship between DM and TB and suggests the need for more comprehensive cohort studies to clarify these associations between TB index cases and their contacts.

Despite strong recommendations in national [11] and international guidelines [7] for screening and testing all contacts of people living with HIV, less than half of these contacts were screened in our study, with only 58.8% (n=18,403) undergoing examination. These findings indicate the urgency of implementing tailored strategies to ensure an inclusive and effective approach in controlling TB and HIV among their contacts. We also observed that compared to contacts of male index cases, contacts of female index cases showed a higher likelihood of being diagnosed with TB, reflecting a pattern observed in other studies in the literature [9].

Our study shows a higher prevalence of TB among contacts of index patients belonging to groups with historical characteristics of social vulnerability, such as those with low education, black and mixed-race ethnicity, homelessness, foreigners, and residents in overcrowded populated areas. These groups, likely due to their precarious housing conditions, restricted access to healthcare services, low income, and unemployment, are more likely to come into contact with patients with active TB, thereby facilitating the spread of the disease among contacts [37,38].

Finally, we found an inverse association between the age of index cases and the prevalence of TB among their contacts, consistent with other studies on the subject [9,39]. This finding is understandable, as children are prioritised in contact tracing policies due to the high likelihood of disease transmission occurring within the family environment [11]. Therefore, identifying index patients under 10 years old might trigger more intensive case-finding efforts within healthcare services [9]. Hence, we suggest that improving contact investigation across all age groups of index patients could have a significant impact on TB prevention and treatment within the state and the country as a whole.

### Limitations and strengths

Our study also had some limitations. The unavailability of data on the characteristics of contacts, including their relationship with the index case and genotypic matching, limited our ability to thoroughly assess whether these factors could influence the likelihood of being examined and the potential determinants of TB among them. This information is not routinely recorded in TBWEB. In addition, we assumed that the lack of examination for some contacts was not related to variables other than those assessed in the index cases. Due to the cross-sectional design of the study, we were unable to establish the time interval between the initial TB report of the index case and the development of the disease among their contacts. This limitation prevents us from determining whether the cases are co-prevalent or incident. However, we believe they are likely co-prevalent, given that the study focused exclusively on new TB cases and employed an investigation algorithm that typically prioritises recent cases to identify active disease outbreaks. Moreover, we cannot confirm whether the contacts acquired TB through direct transmission from the index patient, external exposure, or reactivation of LTBI. These considerations underscore the importance of including such information in future longitudinal studies to better elucidate transmission patterns, disease risk under different circumstances, and the cost-effectiveness of contact investigations. This would provide more robust evidence. Furthermore, such studies could determine whether conducting contact screening for all cases would increase yield compared to current symptom-based recommendations, which focus on individuals with bacteriologically confirmed pulmonary TB, children aged five years or younger, and people living with HIV.

Despite these limitations, the study has several strengths that deserve highlighting. First, the data for this evaluation were routinely collected by the state TB control programme and, therefore, accurately reflect programmatic conditions in low- and middle-income settings with a high TB burden. Consequently, our results are likely generalisable to similar settings where the WHO currently recommends contact investigations. Second, our analysis contributes to a growing body of literature that assesses the effectiveness of TB contact investigation strategies and their performance across different groups and predisposing factors. Third, we employed weighting strategies to ensure the representativeness of the number of contacts screened in the prevalence results, allowing us to estimate the probable number of underreported cases. Finally, the study’s strength lies in the large number of contacts screened and examined compared to previous studies.

In summary, we recommend strengthening and expanding contact investigations for all TB index cases to facilitate early detection and appropriate treatment of new cases. On the other hand, in resource-limited settings, priority should be given to investigating contacts of specific index cases, such as those in socially vulnerable groups, including children and adults, and cases indicating more severe disease. These findings have significant implications for public health policies related to TB control, not only in the state of São Paulo but also in other regions with similar contexts.

## Data availability statement

Due to the ethical reason, data sharing is not applicable.

## Acknowledgements

We would like to thank all staff of the Tuberculosis Control Division at the Epidemiological Surveillance Center "Prof Alexandre Vranjac" of the São Paulo State Department of Health, Brazil.

## Contributions

JMNS and FADQ conceptualized the study. JMNS conducted data collection, organization, and analysis. FADQ provided supervision and contributed to the planning of the analyses. All authors participated in interpreting the results. JMNS drafted the initial version of the manuscript. FADQ made substantial contributions to the revision, and both JMNS and FADQ reviewed and approved the final version.

## Competing interest

The authors declare none.

## Funding

This study was financed in part by the Coordenação de Aperfeiçoamento de Pessoal de Nível Superior - Brazil (CAPES) - Finance Code 001 as a Brazilian CAPES scholarship to JMNS. The funders had no role in the study design, data collection and analysis, decision to publish, or preparation of the manuscript.

## Ethical standard

The study was approved by the Research Ethics Committee of the School of Public Health at the University of São Paulo (protocol number: 4,285,870).

## Supplementary Material

### Supplementary methods: Variables

**Table S1.**
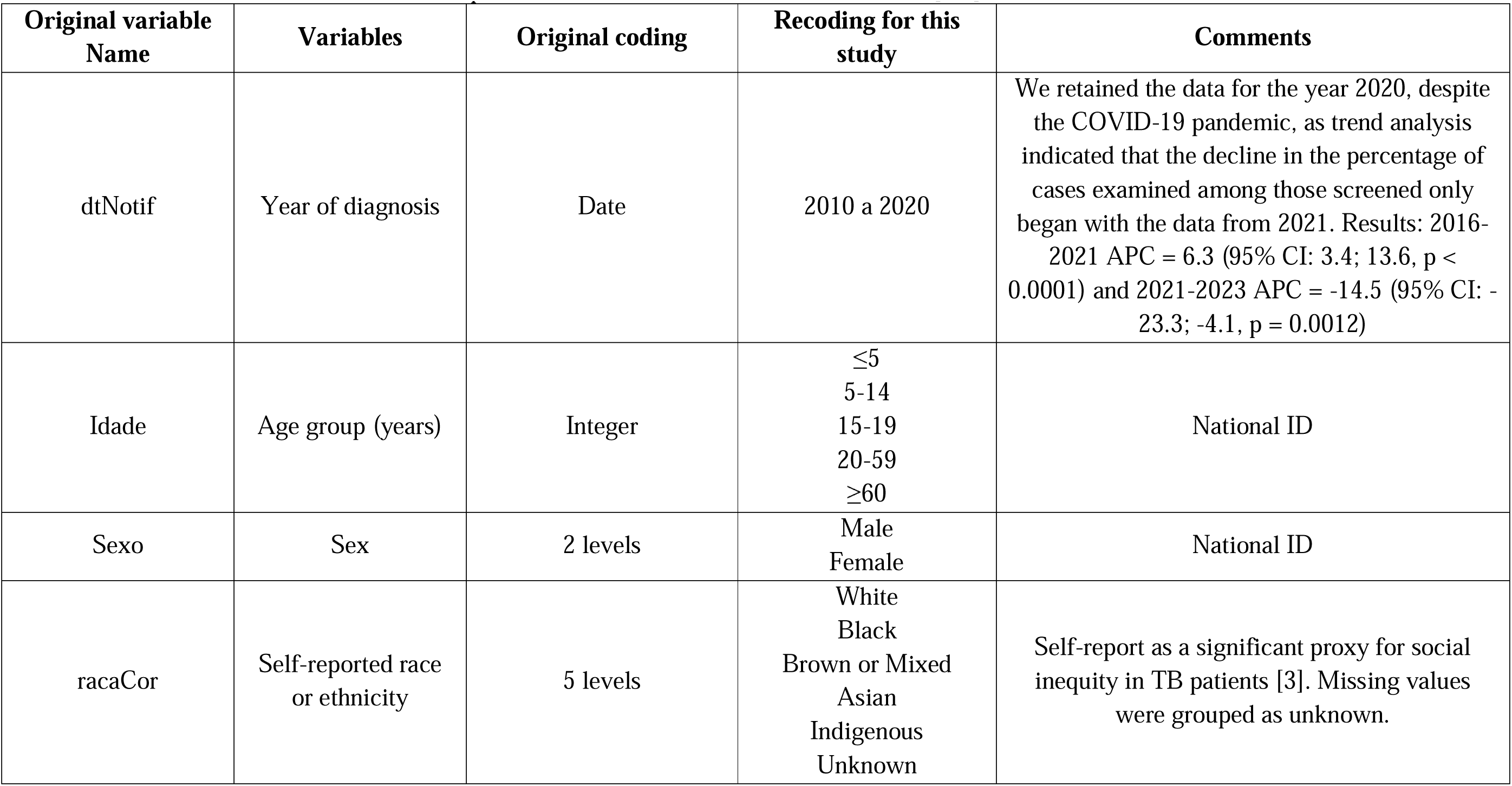
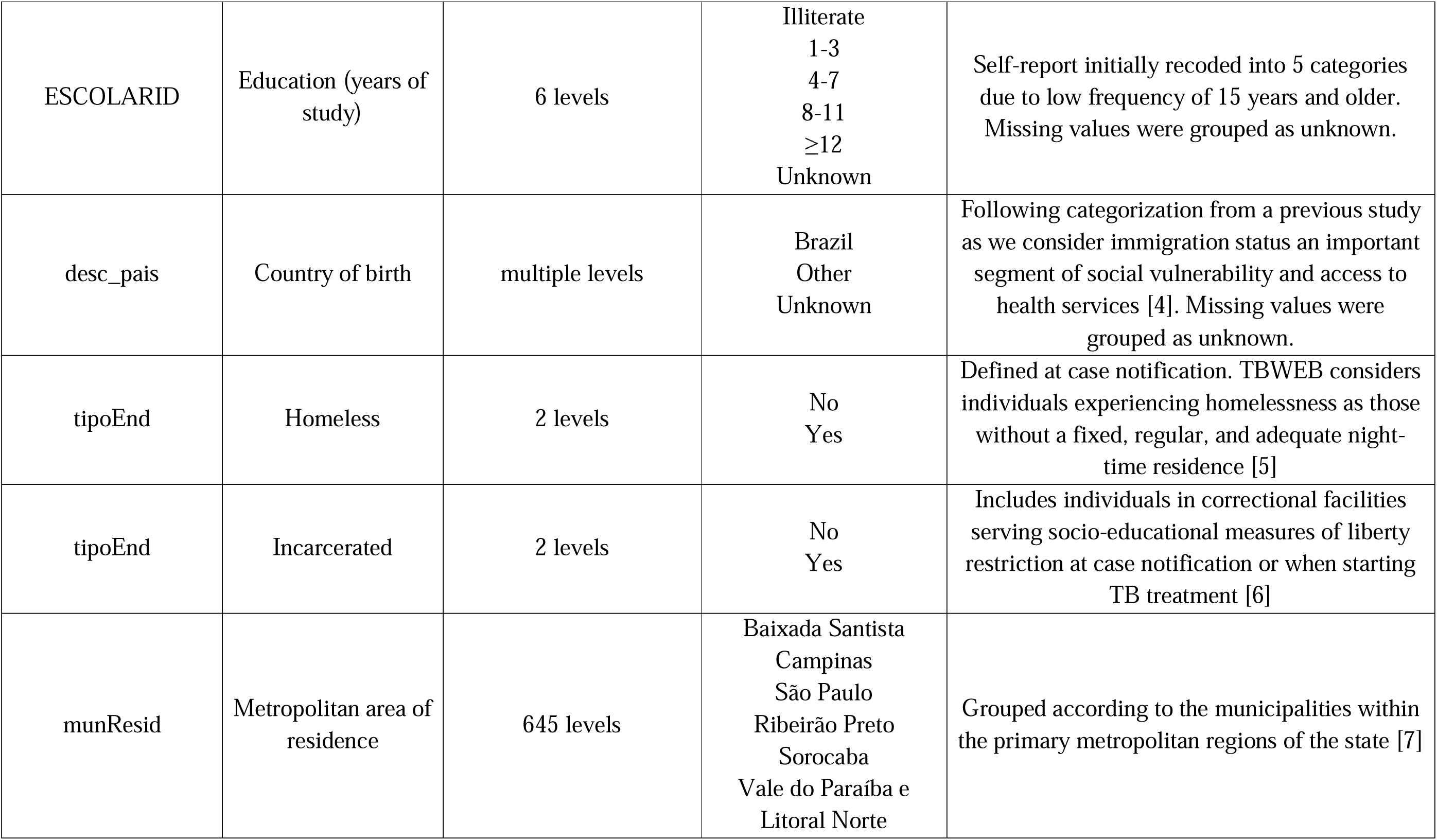
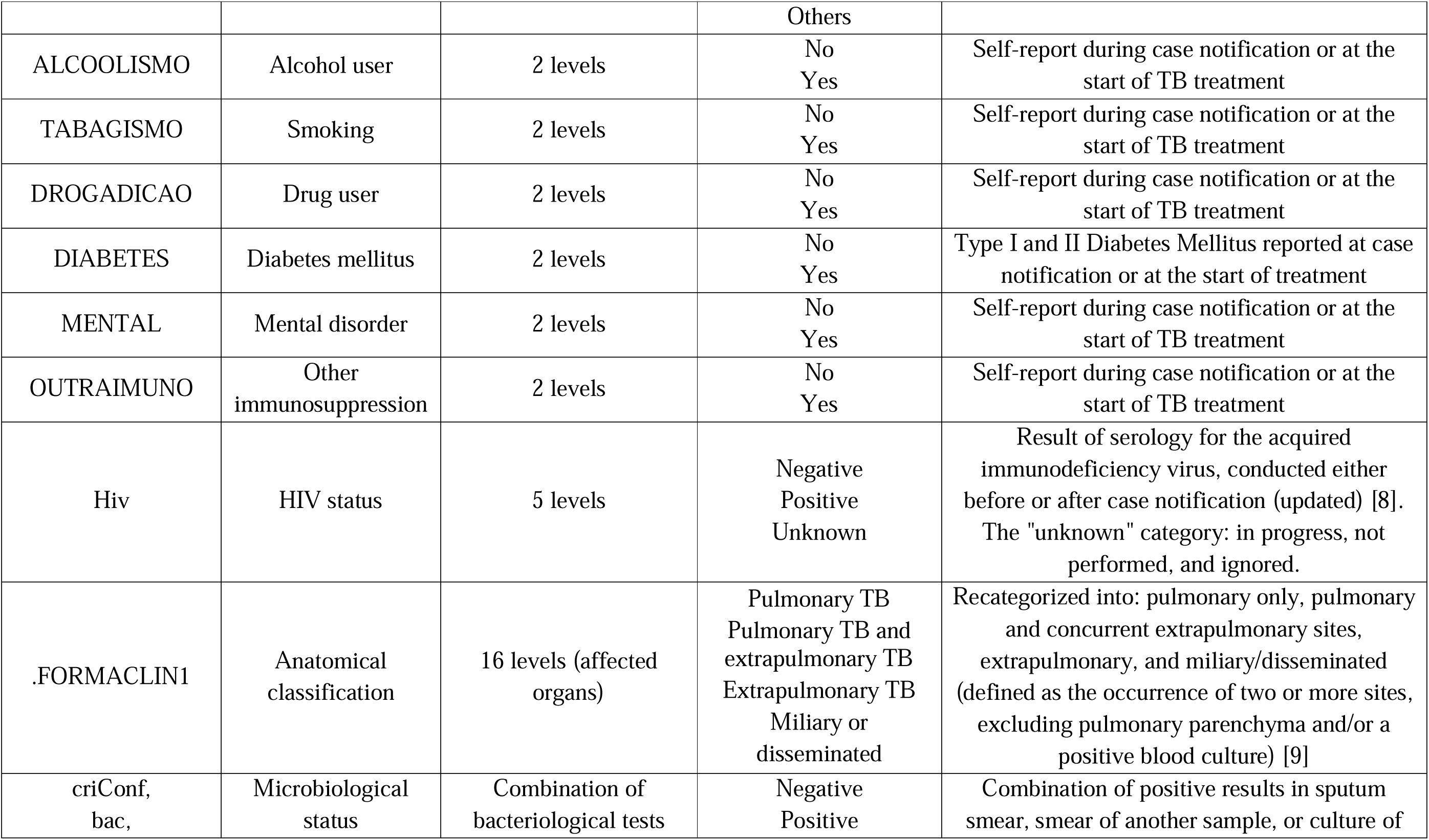
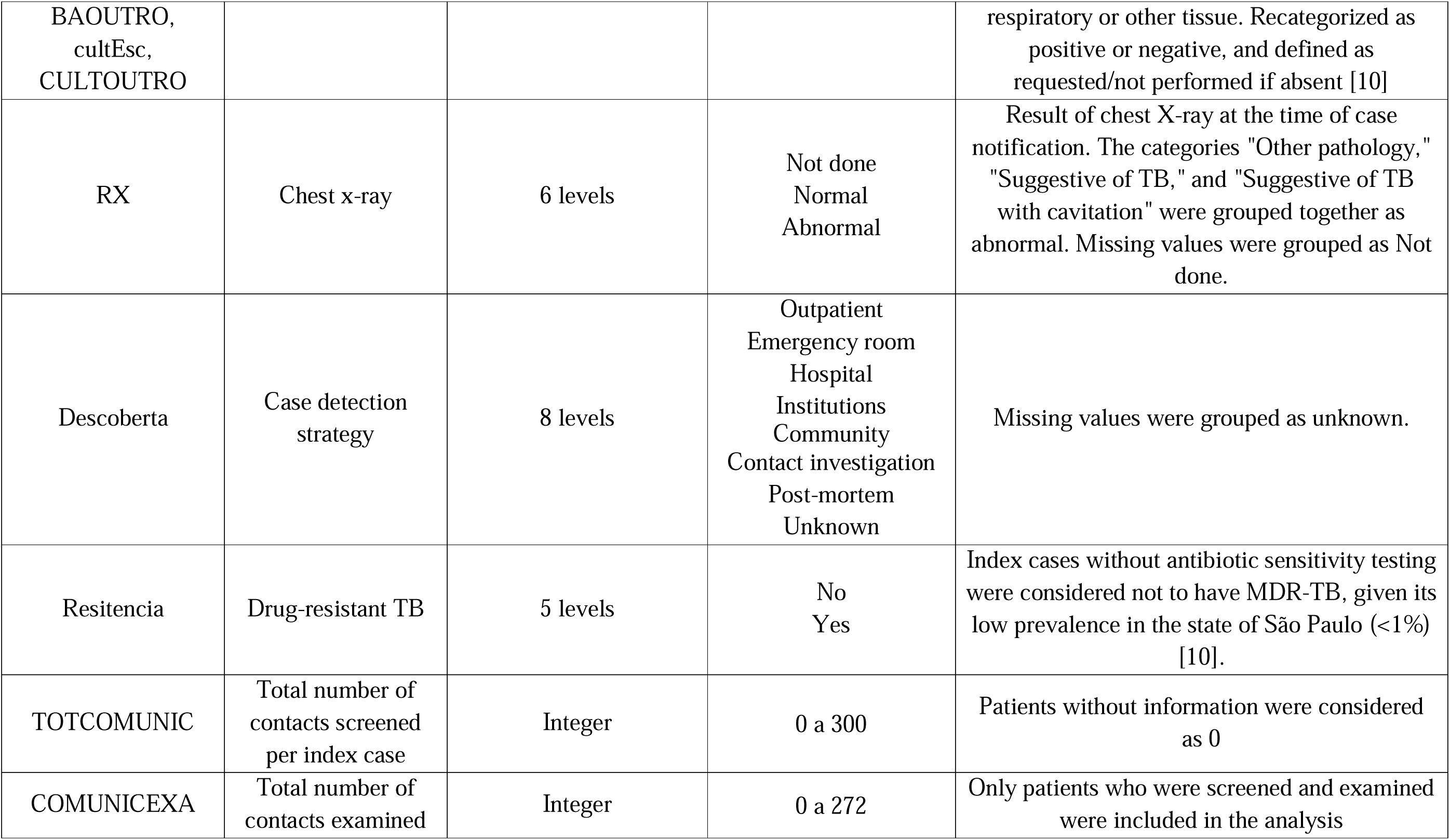
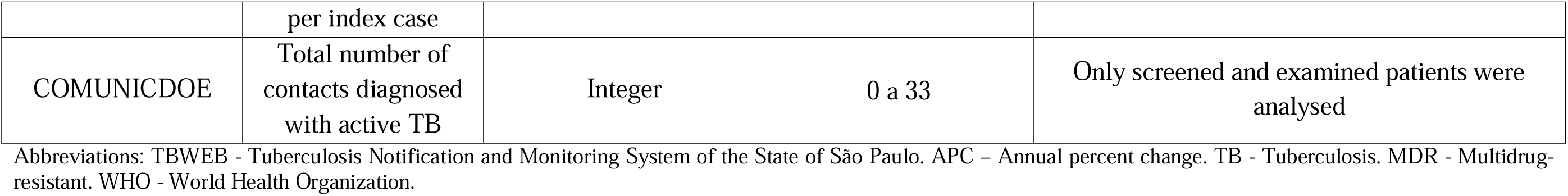
Individual variables related to the patient, and the disease available in TBWEB [1,2].

### Supplementary methods: Contact investigation procedure

**Figure S1.**
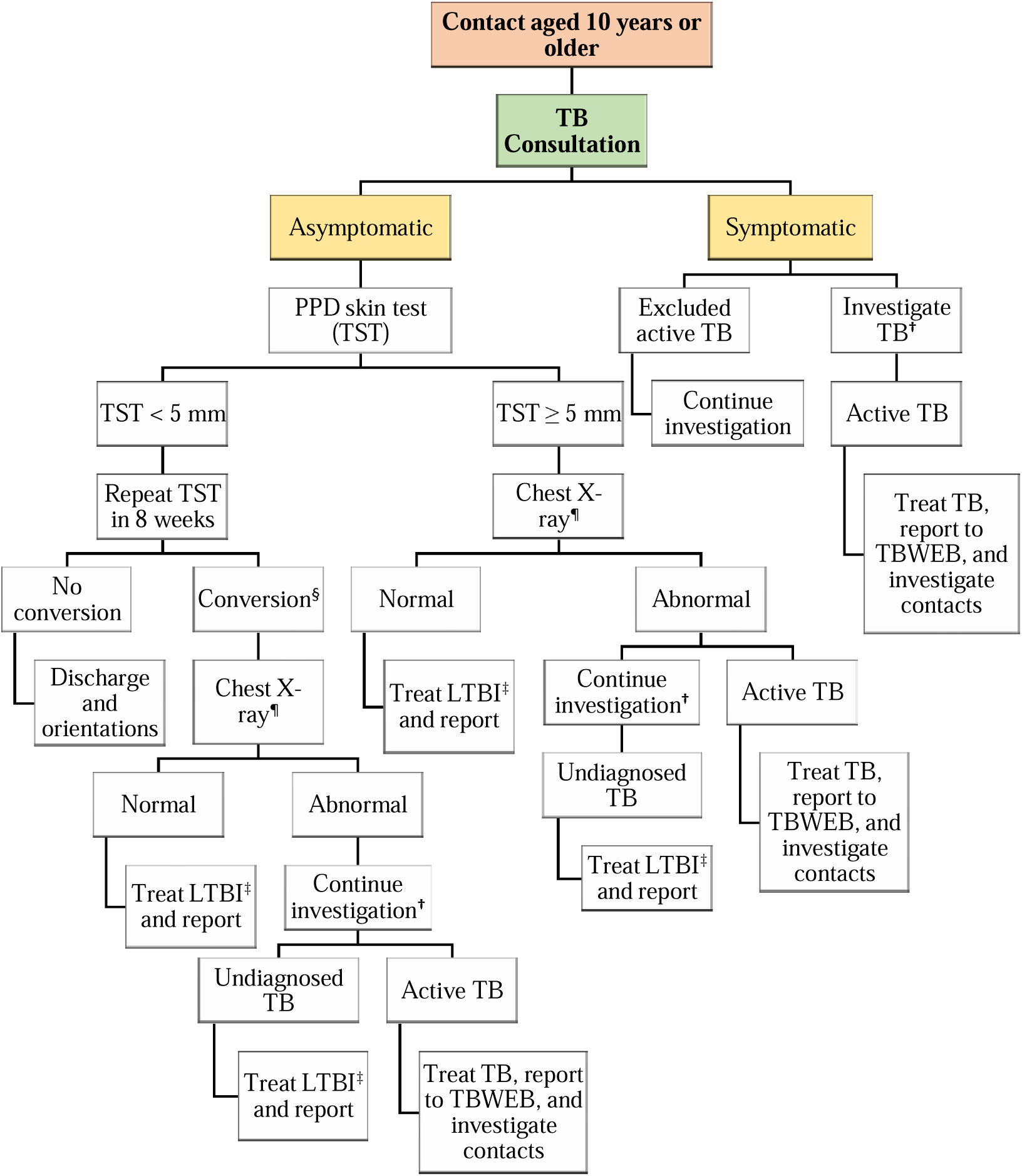
Adapted national algorithm for the investigation of adult and adolescent household contacts (aged 10 years and older) [11,12]. Abbreviations: Abbreviations: TBWEB - Tuberculosis Notification and Monitoring System of the State of São Paulo. TB - Tuberculosis LTBI - Latent tuberculosis infection. TST - Tuberculin skin test. PPD - Purified protein derivative. **^†^**Investigate active TB: clinical assessment, laboratory tests (Xpert MTB/RIF test or smear microscopy, culture, and drug susceptibility testing, when indicated), and chest X-ray. In case of suspected extrapulmonary TB, refer for specific tests at the referral service ^¶^Check for suggestive TB changes on chest X-ray ^‡^In case of HIV-negative pregnant women, treat LTBI only after delivery. ^§^When there is an increase of at least 10 mm compared to the previous TST.

**Figure S2.**
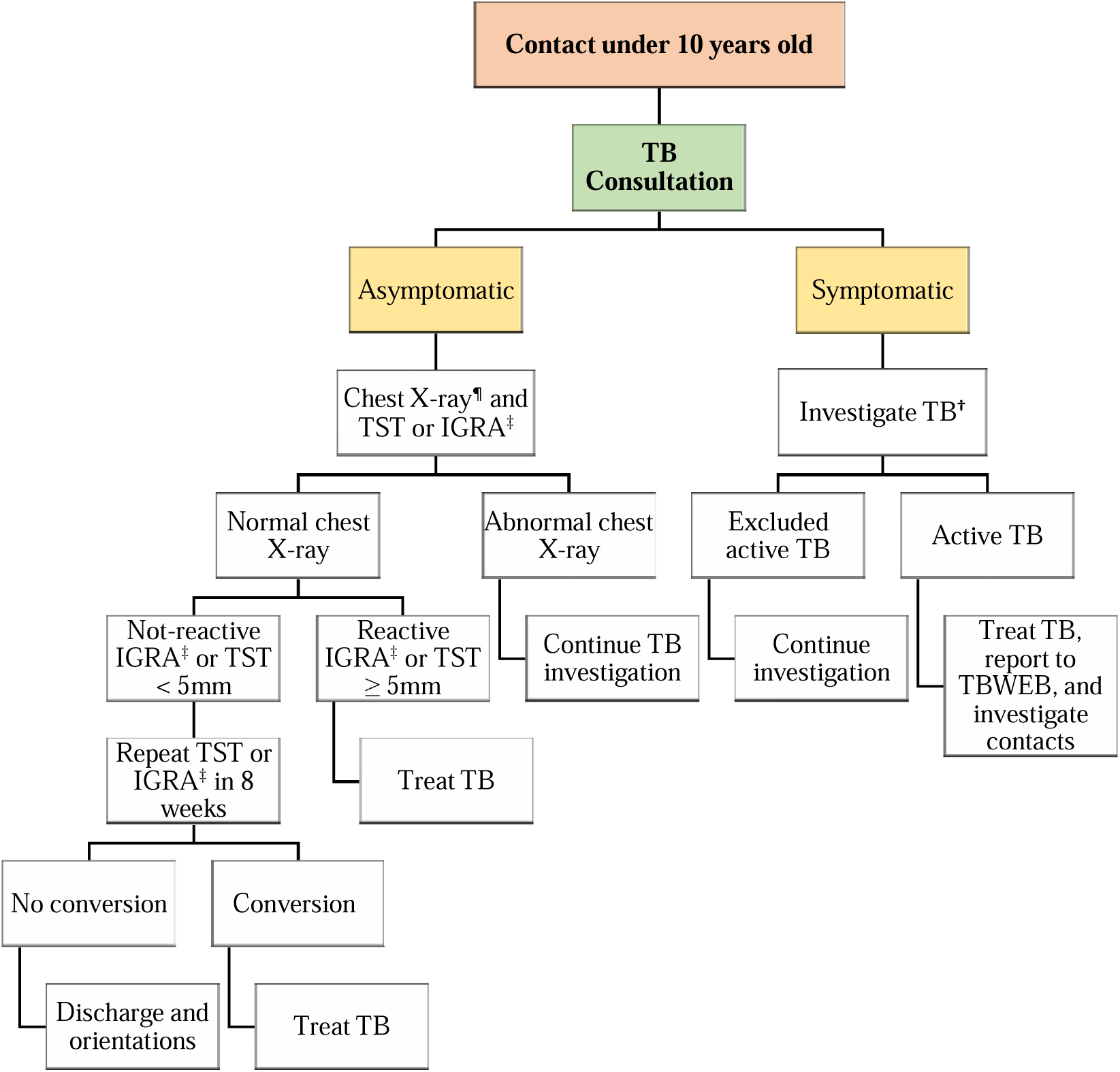
Adapted national algorithm for the investigation of household contacts of children (under 10 years old) [11,12]. Abbreviations: TBWEB - Tuberculosis Notification and Monitoring System of the State of São Paulo. TB – Tuberculosis. TST - Tuberculin skin test. IGRA - Interferon Gamma Release Assay. **^†^**Investigate active TB: clinical assessment, laboratory tests (Xpert MTB/RIF test or smear microscopy, culture, and drug susceptibility testing, when indicated), and chest X-ray. In case of suspected extrapulmonary TB, refer for specific tests at the referral service. ^¶^Check for suggestive TB changes on chest X-ray. ^‡^Perform IGRA only in children aged 2 to 10 years (in children under 2 years and over 10 years, perform TST). ^§^When there is an increase of at least 10 mm compared to the previous PPD TST. It is worth noting that the TST stimulates the immune response to BCG administered at birth, hence the need for this increase in TST after an initial assessment

### Supplementary methods: Predictive model

**Figure S3.**
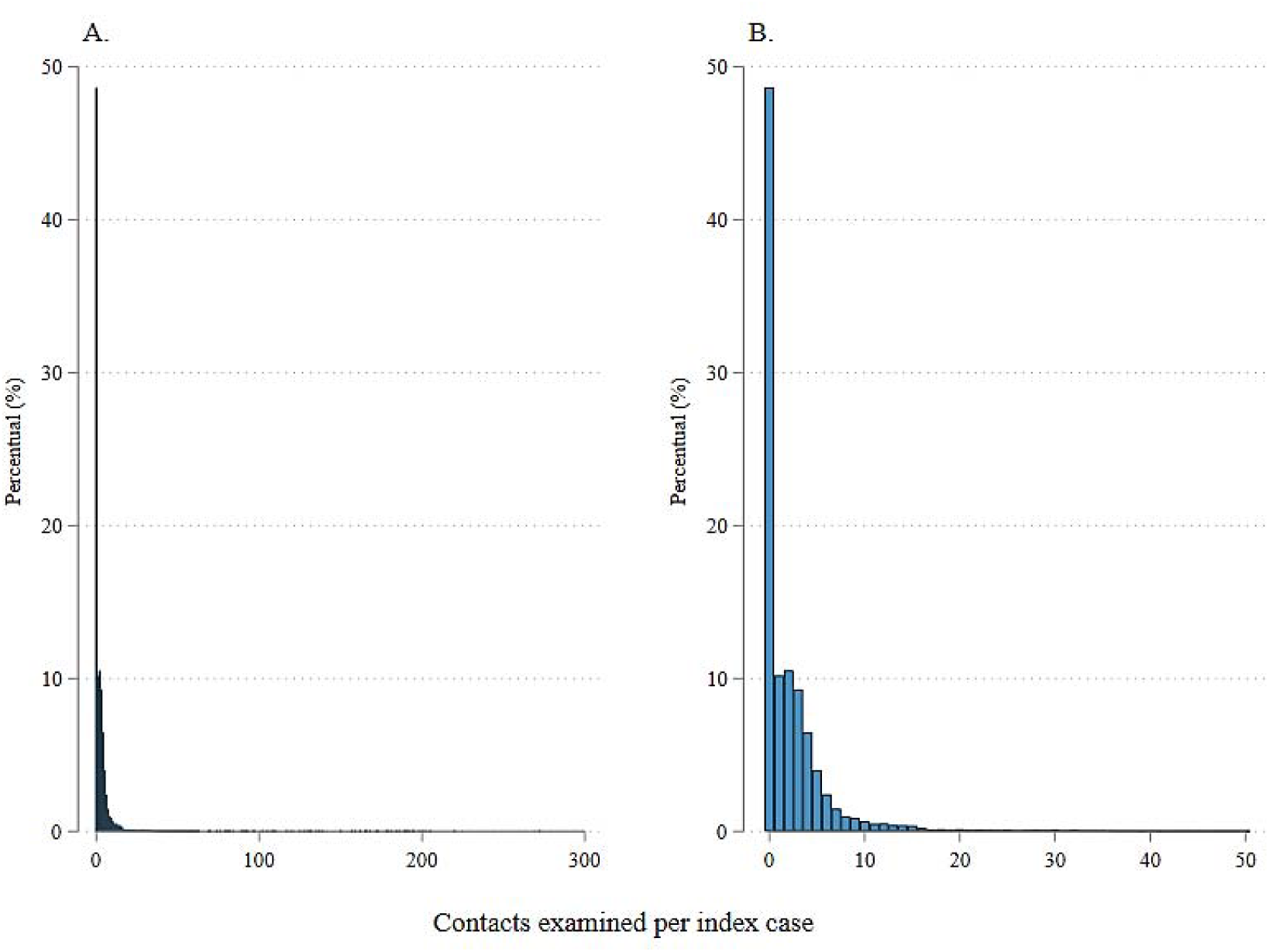
Distribution of the number of contacts examined per tuberculosis index case. A. All contacts. B. Subset of contacts (only 0-50 contacts per index case).

Figure S3 shows the distribution of the dependent variable in the study, the number of contacts examined per index case of tuberculosis (TB). We tested several predictive models, using the number of contacts examined per index case as the dependent variable, patient characteristics as predictor variables, and the total number of contacts screened as an exposure variable. The covariates of interest were: year of diagnosis, sex, age group, race or ethnicity, education, country of birth, homelessness, incarcerated, metropolitan area of residence, case detection strategies, anatomical classification of TB, microbiological status, drug-resistant TB, chest x-ray, HIV serology, and other comorbidities.

Initially, we evaluated the Poisson regression model (PRM) to predict the count of contacts examined among those identified by TB index cases, taking into account the covariates of interest [13]. No multicollinearity was detected among the variables, as all Variance Inflation Factor (VIF) values were below 5, ranging from 1.03 to 1.29. However, the final model did not fit well (Figure S4A). This occurred because the Poisson distribution assumes that the data are equidispersed, meaning the mean equals the variance, which was not observed [14]. The observed variance was approximately 107% greater than the mean (*χ*^2^*person*/df =2,07 e p < 0.001).

Therefore, we opted to use negative binomial regression model (NBRM) instead of PRM, as it allows for modelling the variance more appropriately, reflecting the unobserved heterogeneity in the data [13]. The final model provided a better fit than the PRM (Figure S4B), and the likelihood ratio chi-square test, which checks if the alpha parameter equals zero, resulted in a p-value of less than 0.001, indicating data dispersion (different mean and variance) and making this model more appropriate than the previous one.

However, this observed variability may be attributed to an excessive number of zeros in the data (Figure S3) [15], In the context of the number of contacts examined, this could be characterized by non-registration or, for some reason, the examination not being performed, which seems plausible given that 30.8% of identified contacts were not examined [16]. This translates to a significant number of observed zeros, totalling 36,235, compared to the 23,973 predicted by the model (a ratio of 1.51), within a dataset of 131,055 observations.

To address this situation, zero-inflated models stand out as a viable solution, as they overcome the limitations of previous models by considering not only the dispersion but also the excess zeros. This is achieved by modifying the mean structure, allowing zeros to be generated through two distinct processes: one for null counts (inflated) and another for non-null counts (non-inflated) [13–15]. For the zero-inflated part, the link function of the mean to the linear predictor is given by the logit link, defined as [13]:

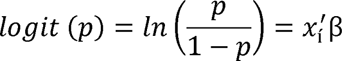

For the non-inflated part, the link function is the same as used in the Poisson model, defined as [13]:

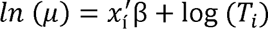

Where p is the probability of the event, x_i_ is the vector of covariates, *β* is a parameter vector (k × 1) of unknown coefficients, and T_i_ is the exposure variable.

Given this context, zero-inflated Poisson (ZIP) and zero-inflated negative binomial (ZINB) regression models were evaluated [15]. However, the latter did not converge, unlike the former. The final ZIP regression model demonstrated superior fit compared to the previous PRM and NBRM (Figures S4C). Thus, the proposed model effectively addressed the issue of excess zeros, estimating that, based on the analysed covariates, approximately 69% of identified contacts would be examined, which is very close to the observed value of 69.2%.

It is important to note that the model selection was based on multiple criteria, including graphical analysis (Figure S4), McFadden’s adjusted coefficient of determination (Pseudo-R²), as well as AIC (Akaike’s Information Criterion) and BIC (Bayesian Information Criterion) criteria [13,14]. The ideal model was the one that presented the best visual fit, a higher pseudo-R² value, and the lowest information criteria scores (Table S2) [17].

**Figure S4.**
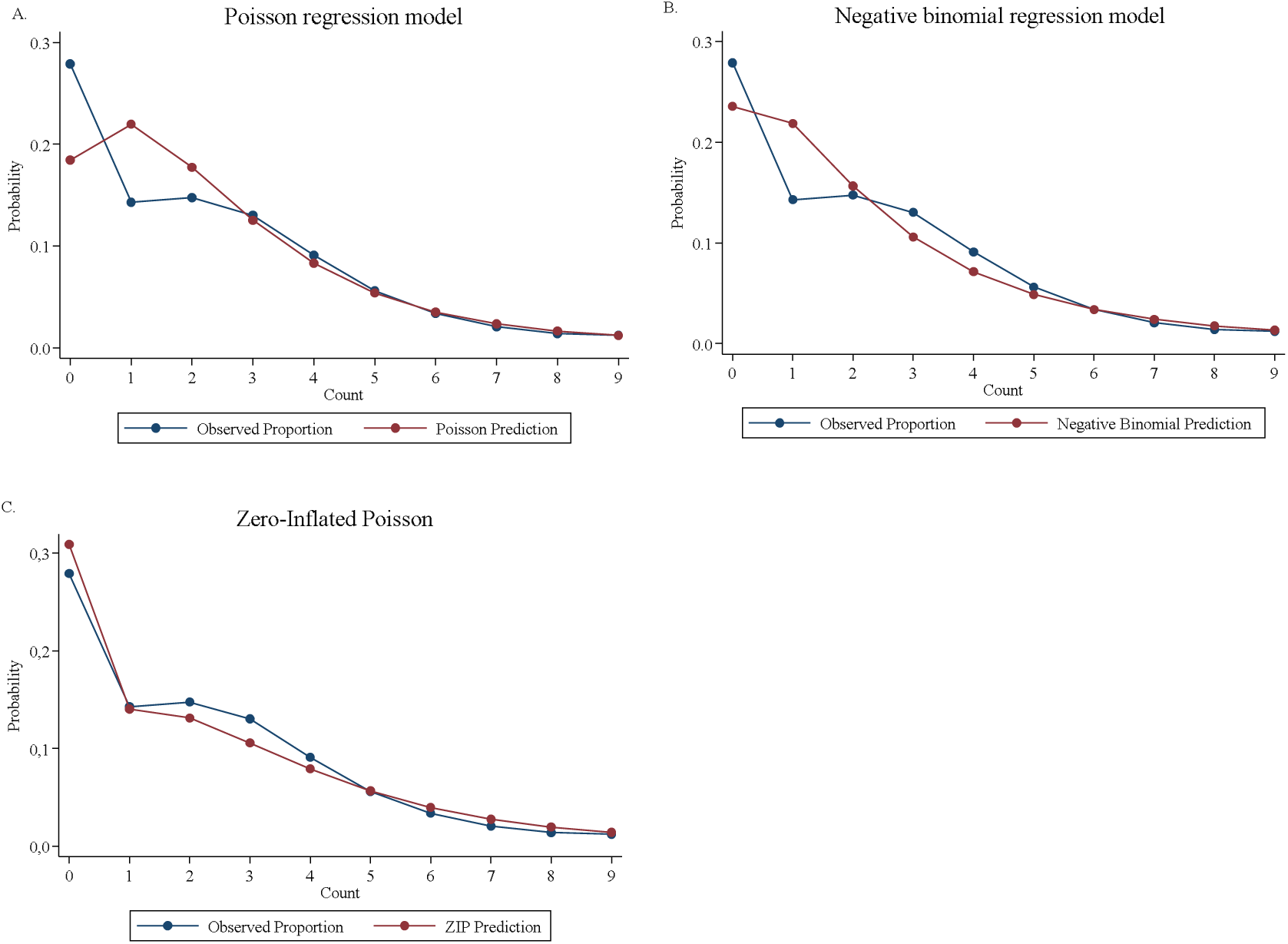
Mean predicted probabilities and observed proportions for each count of contacts examined per index case among count models. A. Poisson regression model. B. Negative binomial regression model. C. Zero-Inflated Poisson.

**Table S2.**
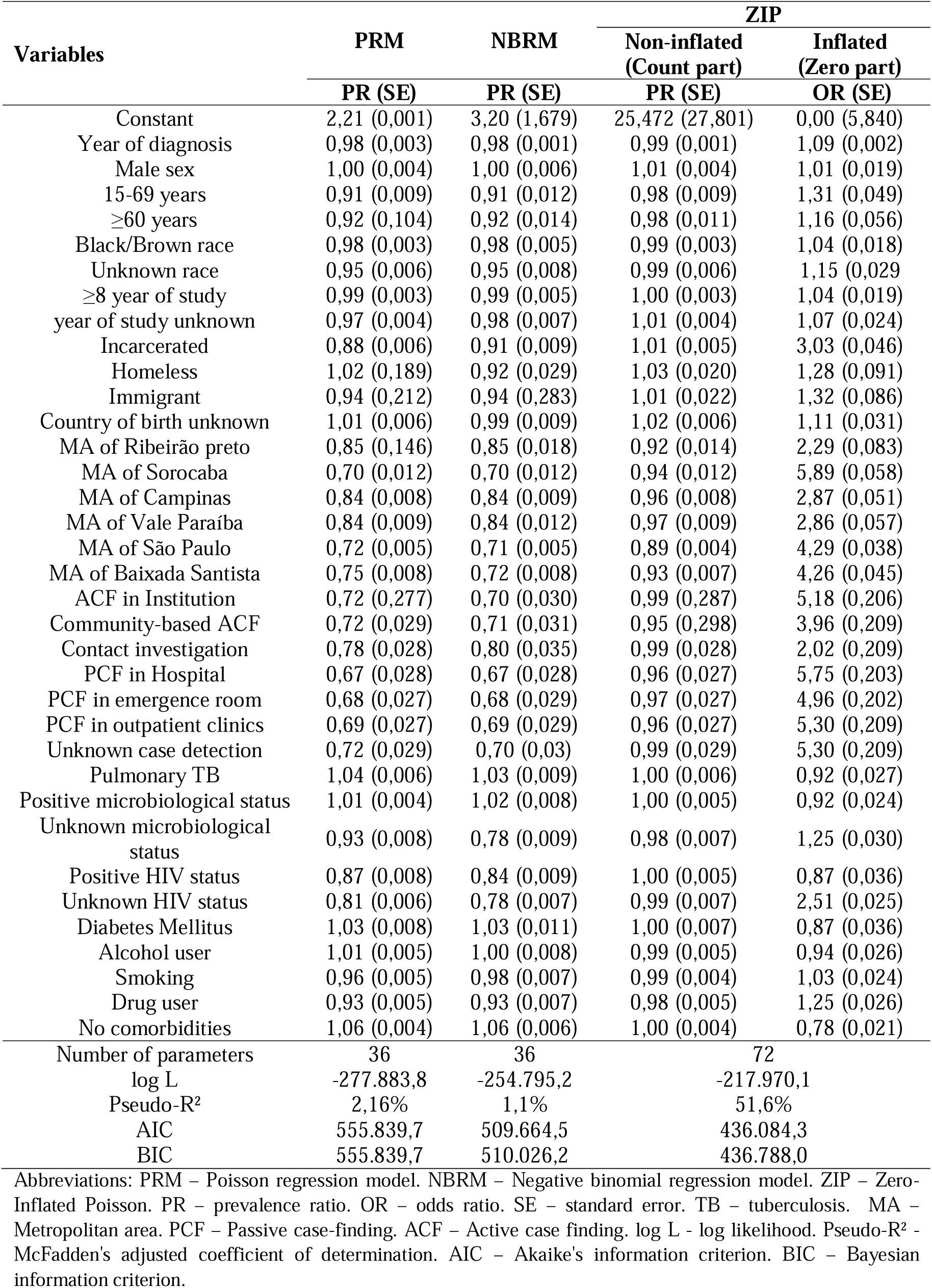
Summary of regression models.

### Supplementary methods: Collider bias

We aimed to evaluate the characteristics of index TB cases among contacts. However, these factors may be associated with the likelihood of contacts being examined, as well as other unknown factors that could influence both this probability and TB itself. Consequently, access to examination becomes a collider, and restricting the analysis to only those contacts who were examined creates an alternative path that can lead to a spurious relationship (collider bias), as shown in the figure below. Therefore, by weighting by the inverse probability of being examined among those screened, we avoid this alternative path and thus minimise the introduction of bias in the association between risk factors and TB.

**Figure S5.**
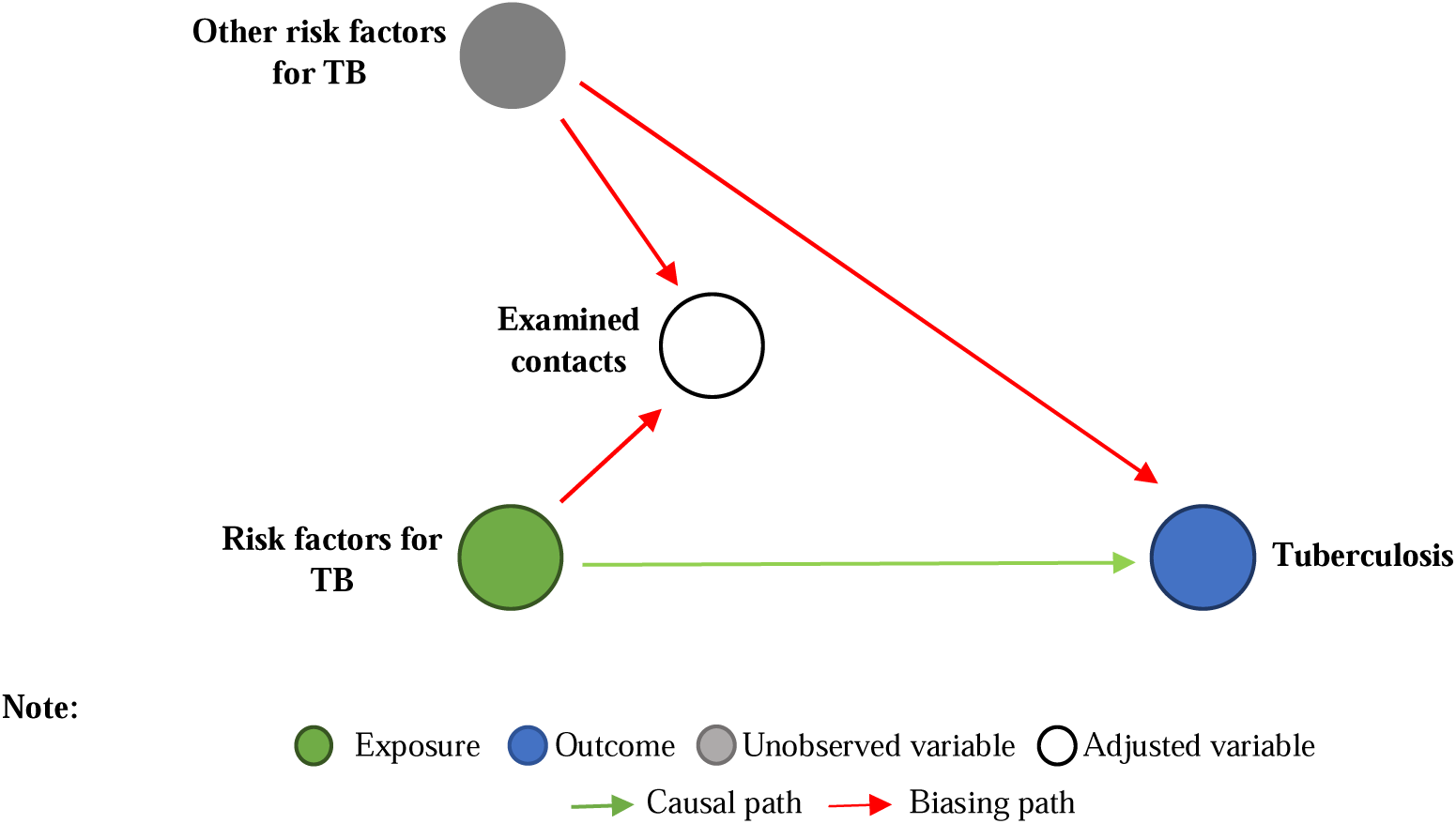
Representation of collider bias when restricting the analysis to examined contacts in the study.

### Supplementary methods: Hierarchical conceptual framework

**Figure S6.**
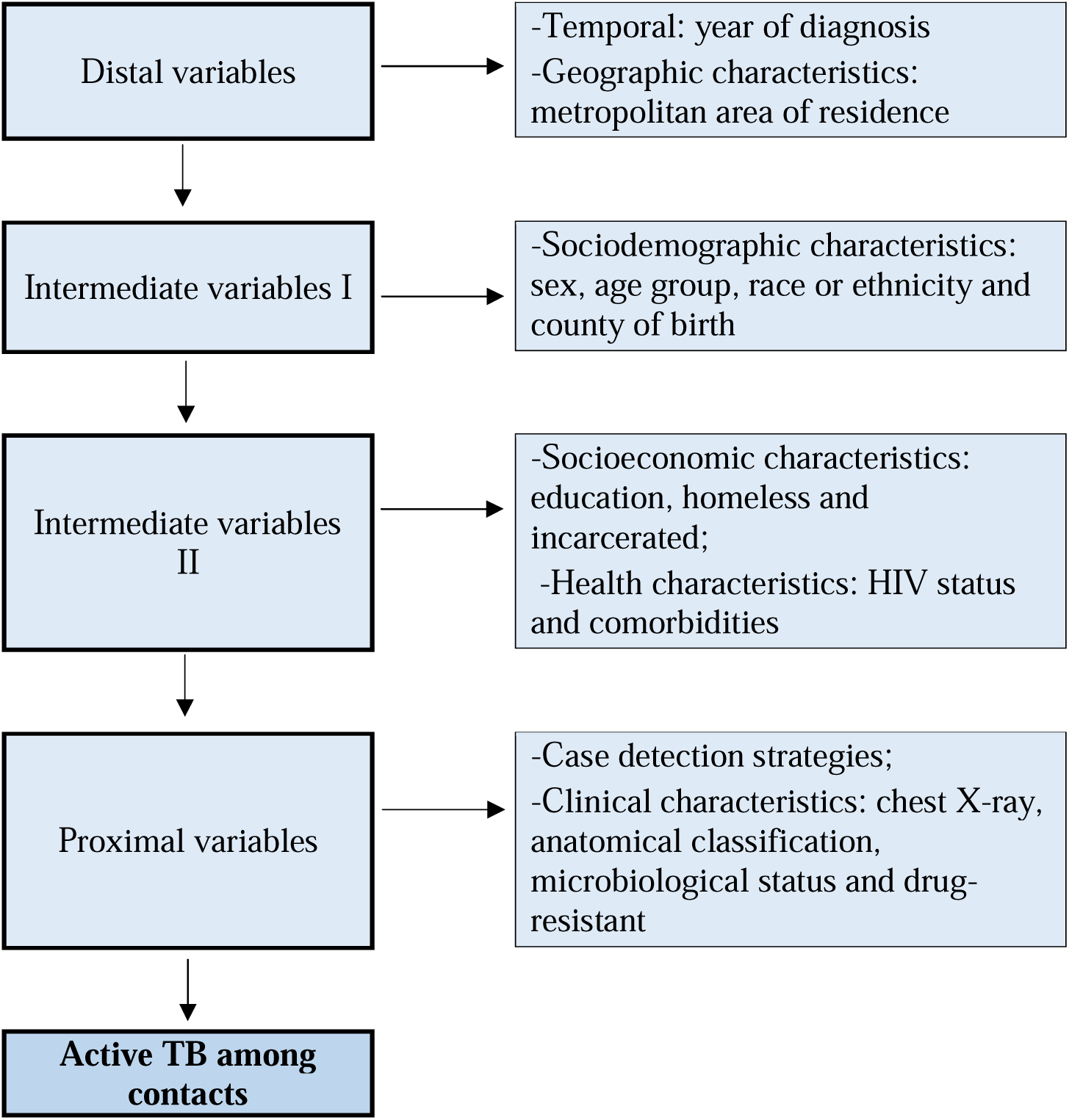
Hierarchical conceptual framework for adjusted multilevel Poisson regression models.

### Supplementary results: Effect of municipalities on TB prevalence

**Table S3.**
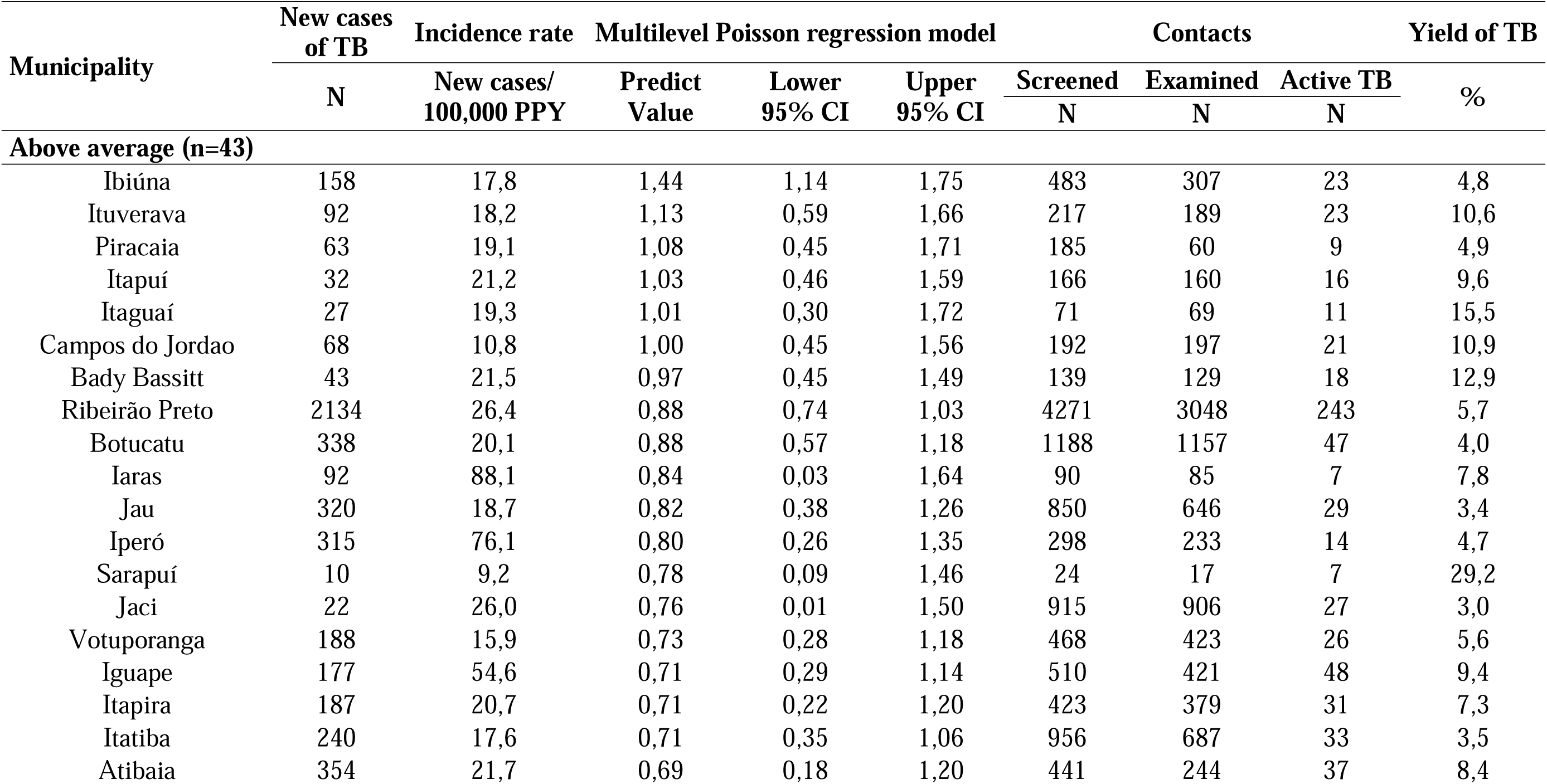
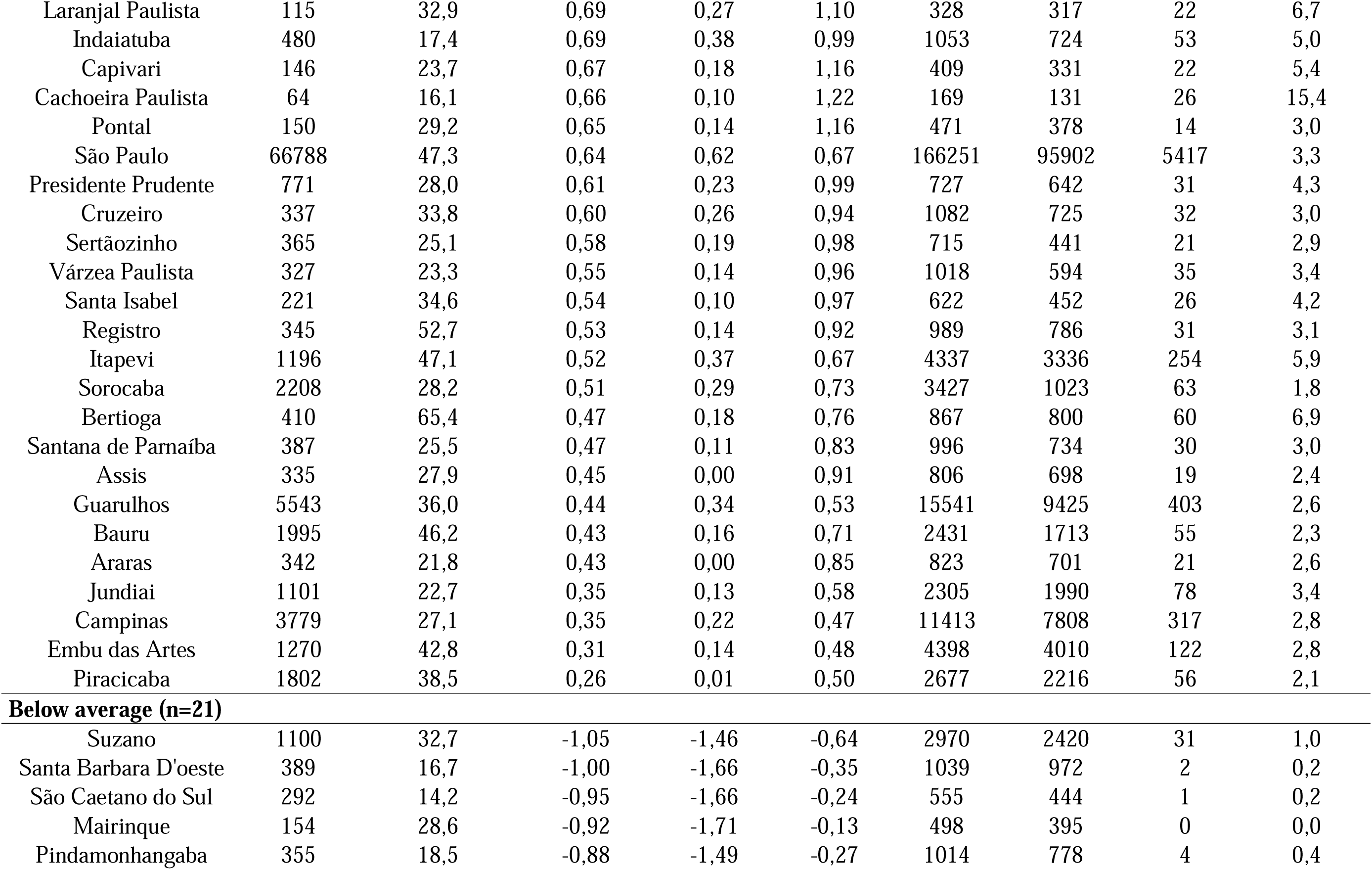
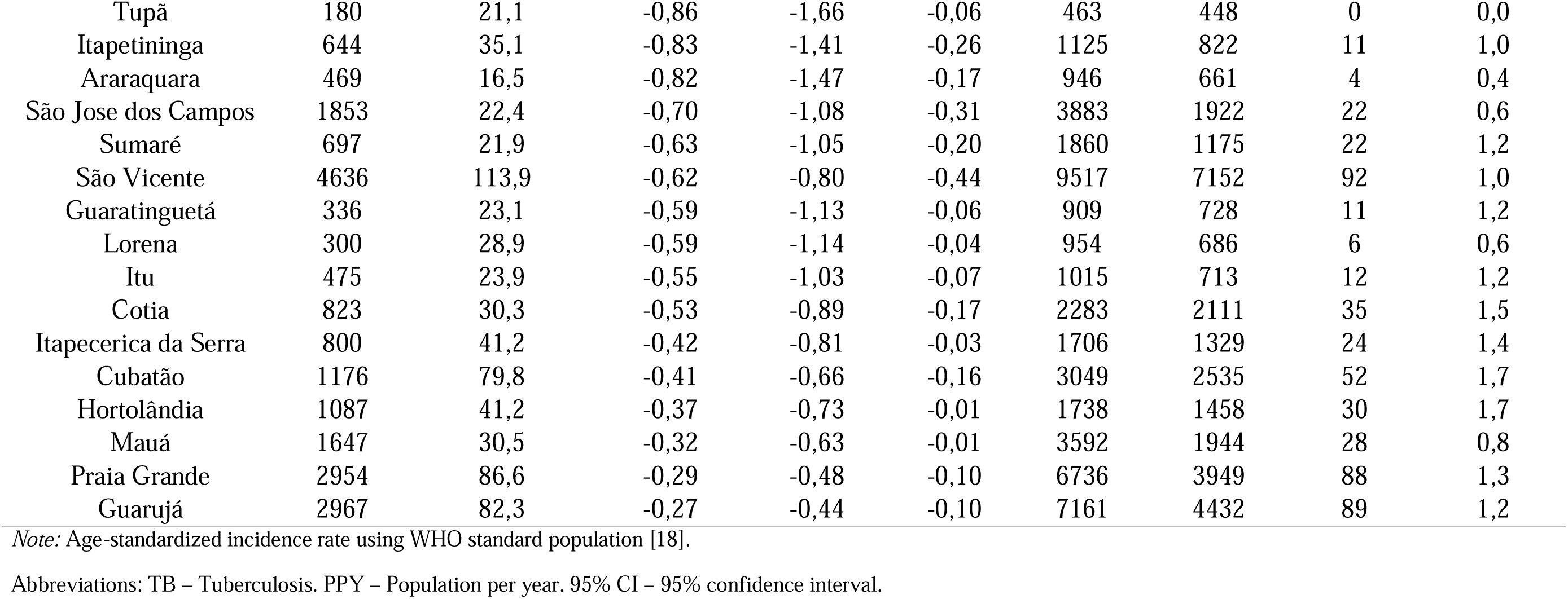
Epidemiological indicators, predict variation, and performance of tuberculosis contact investigation strategy by municipality in the state of São Paulo, Brazil, from 2010 to 2020.

## Notes

### Competing Interest Statement

The authors have declared no competing interest.

### Funding Statement

This study was financed in part by the Coordenação de Aperfeiçoamento de Pessoal de Nivel Superior - Brazil (CAPES) - Finance Code 001 as a Brazilian CAPES scholarship to JMNS.

