## Supplementary Material for "The yield of tuberculosis contact investigation in São Paulo, Brazil: a community-based cross-sectional study"

**List of figures**

[**Figure S1.** Adapted national algorithm for the investigation of adult and adolescent household contacts (aged 10 years and older) [11,12].	7](#_Toc170174944)

[**Figure S2.** Adapted national algorithm for the investigation of household contacts of children (under 10 years old) [11,12].	8](#_Toc170174945)

**List of tables**

[**Table S1.** Individual variables related to the patient, and the disease available in TBWEB [1,2].	3](#_Toc170063698)

### Supplementary methods: Variables

Table S1. Individual variables related to the patient, and the disease available in TBWEB [1,2].

| **Original variable**  **Name** | **Variables** | **Original coding** | **Recoding for this study** | **Comments** |
| --- | --- | --- | --- | --- |
| dtNotif | Year of diagnosis | Date | 2010 a 2020 | We retained the data for the year 2020, despite the COVID-19 pandemic, as trend analysis indicated that the decline in the percentage of cases examined among those screened only began with the data from 2021. Results: 2016-2021 APC = 6.3 (95% CI: 3.4; 13.6, p < 0.0001) and 2021-2023 APC = -14.5 (95% CI: -23.3; -4.1, p = 0.0012) |
| Idade | Age group (years) | Integer | ≤5  5-14  15-19  20-59  ≥60 | National ID |
| Sexo | Sex | 2 levels | Male Female | National ID |
| racaCor | Self-reported race or ethnicity | 5 levels | White  Black  Brown or Mixed  Asian  Indigenous  Unknown | Self-report as a significant proxy for social inequity in TB patients [3]. Missing values were grouped as unknown. |
| ESCOLARID | Education (years of study) | 6 levels | Illiterate  1-3 4-7 8-11 ≥12  Unknown | Self-report initially recoded into 5 categories due to low frequency of 15 years and older. Missing values were grouped as unknown. |
| desc_pais | Country of birth | multiple levels | Brazil  Other  Unknown | Following categorization from a previous study as we consider immigration status an important segment of social vulnerability and access to health services [4]. Missing values were grouped as unknown. |
| tipoEnd | Homeless | 2 levels | No  Yes | Defined at case notification. TBWEB considers individuals experiencing homelessness as those without a fixed, regular, and adequate night-time residence [5] |
| tipoEnd | Incarcerated | 2 levels | No  Yes | Includes individuals in correctional facilities serving socio-educational measures of liberty restriction at case notification or when starting TB treatment [6] |
| munResid | Metropolitan area of residence | 645 levels | Baixada Santista  Campinas  São Paulo  Ribeirão Preto  Sorocaba  Vale do Paraíba e Litoral Norte  Others | Grouped according to the municipalities within the primary metropolitan regions of the state [7] |
| ALCOOLISMO | Alcohol user | 2 levels | No  Yes | Self-report during case notification or at the start of TB treatment |
| TABAGISMO | Smoking | 2 levels | No  Yes | Self-report during case notification or at the start of TB treatment |
| DROGADICAO | Drug user | 2 levels | No  Yes | Self-report during case notification or at the start of TB treatment |
| DIABETES | Diabetes mellitus | 2 levels | No  Yes | Type I and II Diabetes Mellitus reported at case notification or at the start of treatment |
| MENTAL | Mental disorder | 2 levels | No  Yes | Self-report during case notification or at the start of TB treatment |
| OUTRAIMUNO | Other immunosuppression | 2 levels | No  Yes | Self-report during case notification or at the start of TB treatment |
| Hiv | HIV status | 5 levels | Negative Positive Unknown | Result of serology for the acquired immunodeficiency virus, conducted either before or after case notification (updated) [8]. The "unknown" category: in progress, not performed, and ignored. |
| .FORMACLIN1 | Anatomical classification | 16 levels (affected  organs) | Pulmonary TB  Pulmonary TB and  extrapulmonary TB  Extrapulmonary TB  Miliary or disseminated | Recategorized into: pulmonary only, pulmonary and concurrent extrapulmonary sites, extrapulmonary, and miliary/disseminated (defined as the occurrence of two or more sites, excluding pulmonary parenchyma and/or a positive blood culture) [9] |
| criConf,  bac,  BAOUTRO,  cultEsc,  CULTOUTRO | Microbiological status | Combination of bacteriological tests | Negative Positive | Combination of positive results in sputum smear, smear of another sample, or culture of respiratory or other tissue. Recategorized as positive or negative, and defined as requested/not performed if absent [10] |
| RX | Chest x-ray | 6 levels | Not done  Normal  Abnormal | Result of chest X-ray at the time of case notification. The categories "Other pathology," "Suggestive of TB," and "Suggestive of TB with cavitation" were grouped together as abnormal. Missing values were grouped as Not done. |
| Descoberta | Case detection strategy | 8 levels | Outpatient  Emergency room  Hospital  Institutions  Community  Contact investigation  Post-mortem  Unknown | Missing values were grouped as unknown. |
| Resitencia | Drug-resistant TB | 5 levels | No  Yes | Index cases without antibiotic sensitivity testing were considered not to have MDR-TB, given its low prevalence in the state of São Paulo (<1%) [10]. |
| TOTCOMUNIC | Total number of contacts screened per index case | Integer | 0 a 300 | Patients without information were considered as 0 |
| COMUNICEXA | Total number of contacts examined per index case | Integer | 0 a 272 | Only patients who were screened and examined were included in the analysis |
| COMUNICDOE | Total number of contacts diagnosed with active TB | Integer | 0 a 33 | Only screened and examined patients were analysed |

Abbreviations: TBWEB - Tuberculosis Notification and Monitoring System of the State of São Paulo. APC – Annual percent change. TB - Tuberculosis. MDR - Multidrug-resistant. WHO - World Health Organization.

### Supplementary methods: Contact investigation procedure

Contact aged 10 years or older

TB Consultation

Asymptomatic

PPD skin test (TST)

TST < 5 mm

Repeat TST in 8 weeks

No conversion

Discharge and orientations

Conversion§

Chest X-ray¶

Normal

Treat LTBI‡ and report

Abnormal

Continue investigation†

Undiagnosed TB

Treat LTBI‡ and report

Active TB

Treat TB, report to TBWEB, and investigate contacts

TST ≥ 5 mm

Chest X-ray¶

Normal

Treat LTBI‡ and report

Abnormal

Continue investigation†

Undiagnosed TB

Treat LTBI‡ and report

Active TB

Treat TB, report to TBWEB, and investigate contacts

Symptomatic

Excluded active TB

Continue investigation

Investigate TB†

Active TB

Treat TB, report to TBWEB, and investigate contacts

Figure S1. Adapted national algorithm for the investigation of adult and adolescent household contacts (aged 10 years and older) [11,12].

Abbreviations: Abbreviations: TBWEB - Tuberculosis Notification and Monitoring System of the State of São Paulo. TB - Tuberculosis LTBI - Latent tuberculosis infection. TST - Tuberculin skin test. PPD - Purified protein derivative.

**^†^**Investigate active TB: clinical assessment, laboratory tests (Xpert MTB/RIF test or smear microscopy, culture, and drug susceptibility testing, when indicated), and chest X-ray. In case of suspected extrapulmonary TB, refer for specific tests at the referral service

^¶^Check for suggestive TB changes on chest X-ray

^‡^In case of HIV-negative pregnant women, treat LTBI only after delivery.

^§^When there is an increase of at least 10 mm compared to the previous TST.

Contact under 10 years old

TB Consultation

Asymptomatic

Chest X-ray¶ and TST or IGRA‡

Normal chest X-ray

Not-reactive IGRA‡ or TST < 5mm

Repeat TST or IGRA‡ in 8 weeks

No conversion

Discharge and orientations

Conversion§

Treat TB

Reactive IGRA‡ or TST ≥ 5mm

Treat TB

Abnormal chest X-ray

Continue TB investigation

Symptomatic

Investigate TB†

Excluded active TB

Continue investigation

Active TB

Treat TB, report to TBWEB, and investigate contacts

**Figure S2.** Adapted national algorithm for the investigation of household contacts of children (under 10 years old) [11,12].

Abbreviations: TBWEB - Tuberculosis Notification and Monitoring System of the State of São Paulo. TB – Tuberculosis. TST - Tuberculin skin test. IGRA - Interferon Gamma Release Assay.

**^†^**Investigate active TB: clinical assessment, laboratory tests (Xpert MTB/RIF test or smear microscopy, culture, and drug susceptibility testing, when indicated), and chest X-ray. In case of suspected extrapulmonary TB, refer for specific tests at the referral service.

^¶^Check for suggestive TB changes on chest X-ray.

^‡^Perform IGRA only in children aged 2 to 10 years (in children under 2 years and over 10 years, perform TST).

^§^When there is an increase of at least 10 mm compared to the previous PPD TST. It is worth noting that the TST stimulates the immune response to BCG administered at birth, hence the need for this increase in TST after an initial assessment.

### Supplementary methods: Predictive model

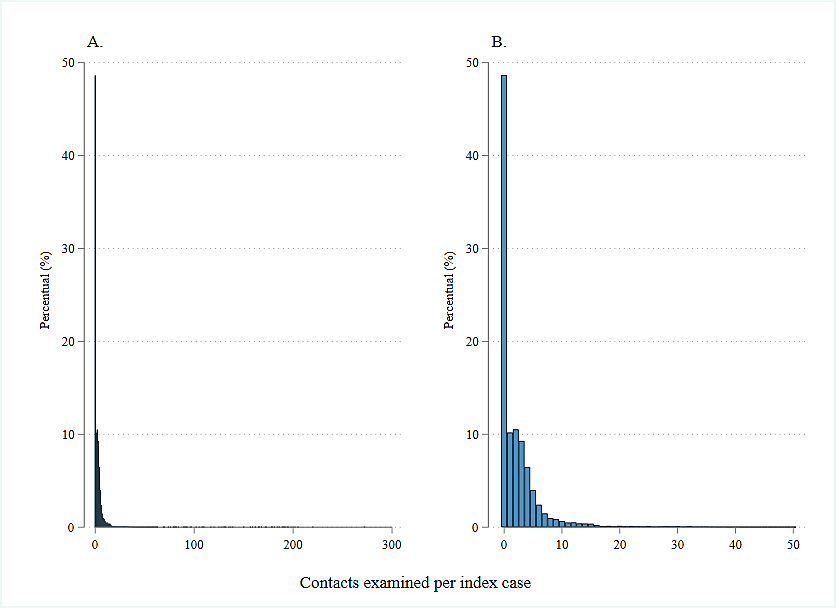

**Figure S3.** Distribution of the number of contacts examined per tuberculosis index case. A. All contacts. B. Subset of contacts (only 0-50 contacts per index case).

Figure S3 shows the distribution of the dependent variable in the study, the number of contacts examined per index case of tuberculosis (TB). We tested several predictive models, using the number of contacts examined per index case as the dependent variable, patient characteristics as predictor variables, and the total number of contacts screened as an exposure variable. The covariates of interest were: year of diagnosis, sex, age group, race or ethnicity, education, country of birth, homelessness, incarcerated, metropolitan area of residence, case detection strategies, anatomical classification of TB, microbiological status, drug-resistant TB, chest x-ray, HIV serology, and other comorbidities.

$$logit \left( p \right)=ln\left( \frac{p}{1-p} \right)=x_{í}^{'}\beta$$

For the non-inflated part, the link function is the same as used in the Poisson model, defined as [13]:

$$ln \left( \mu\right)=x_{í}^{'}\beta+log(T_{i})$$

Where p is the probability of the event, x_i_ is the vector of covariates, *β* is a parameter vector (k × 1) of unknown coefficients, and T_i_ is the exposure variable.

Given this context, zero-inflated Poisson (ZIP) and zero-inflated negative binomial (ZINB) regression models were evaluated [15]. However, the latter did not converge, unlike the former. The final ZIP regression model demonstrated superior fit compared to the previous PRM and NBRM (Figures S4C). Thus, the proposed model effectively addressed the issue of excess zeros, estimating that, based on the analysed covariates, approximately 69% of identified contacts would be examined, which is very close to the observed value of 69.2%.

It is important to note that the model selection was based on multiple criteria, including graphical analysis (Figure S4), McFadden's adjusted coefficient of determination (Pseudo-R²), as well as AIC (Akaike's Information Criterion) and BIC (Bayesian Information Criterion) criteria [13,14]. The ideal model was the one that presented the best visual fit, a higher pseudo-R² value, and the lowest information criteria scores (Table S2) [17].

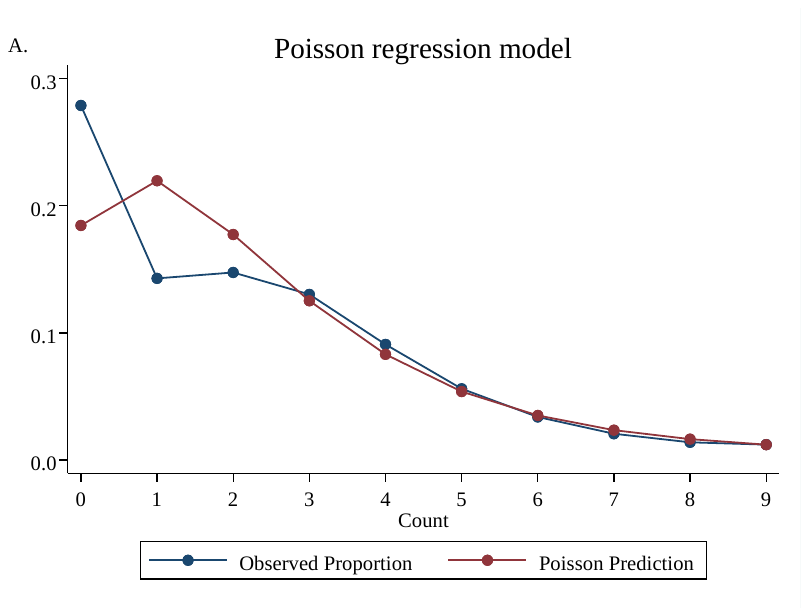

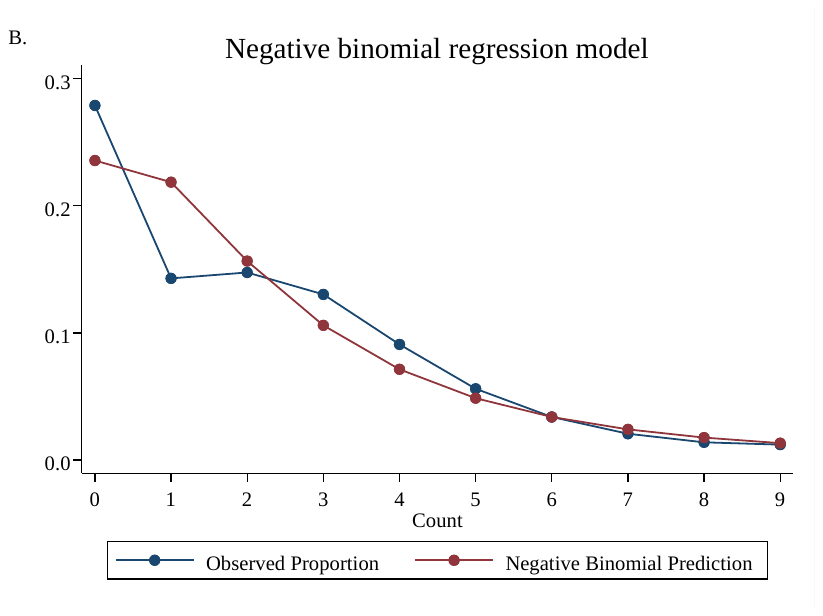

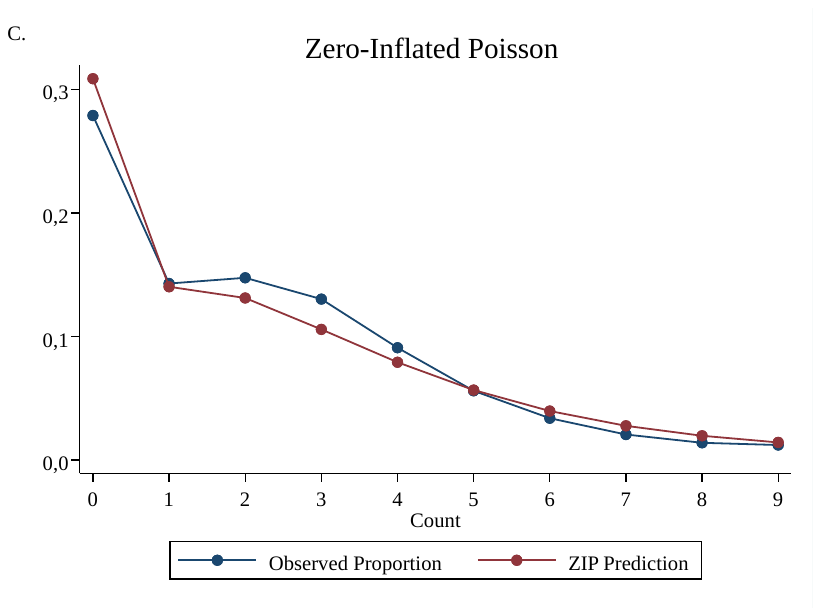

**Figure S4**. Mean predicted probabilities and observed proportions for each count of contacts examined per index case among count models. A. Poisson regression model. B. Negative binomial regression model. C. Zero-Inflated Poisson.

**Table S2.** Summary of regression models.

| **Variables** | **PRM** | **NBRM** | **ZIP** | |
| --- | --- | --- | --- | --- |
|  |  |  | **Non-inflated**  **(Count part)** | **Inflated**  **(Zero part)** |
|  | **PR (SE)** | **PR (SE)** | **PR (SE)** | **OR (SE)** |
| Constant | 2,21 (0,001) | 3,20 (1,679) | 25,472 (27,801) | 0,00 (5,840) |
| Year of diagnosis | 0,98 (0,003) | 0,98 (0,001) | 0,99 (0,001) | 1,09 (0,002) |
| Male sex | 1,00 (0,004) | 1,00 (0,006) | 1,01 (0,004) | 1,01 (0,019) |
| 15-69 years | 0,91 (0,009) | 0,91 (0,012) | 0,98 (0,009) | 1,31 (0,049) |
| ≥60 years | 0,92 (0,104) | 0,92 (0,014) | 0,98 (0,011) | 1,16 (0,056) |
| Black/Brown race | 0,98 (0,003) | 0,98 (0,005) | 0,99 (0,003) | 1,04 (0,018) |
| Unknown race | 0,95 (0,006) | 0,95 (0,008) | 0,99 (0,006) | 1,15 (0,029 |
| ≥8 year of study | 0,99 (0,003) | 0,99 (0,005) | 1,00 (0,003) | 1,04 (0,019) |
| year of study unknown | 0,97 (0,004) | 0,98 (0,007) | 1,01 (0,004) | 1,07 (0,024) |
| Incarcerated | 0,88 (0,006) | 0,91 (0,009) | 1,01 (0,005) | 3,03 (0,046) |
| Homeless | 1,02 (0,189) | 0,92 (0,029) | 1,03 (0,020) | 1,28 (0,091) |
| Immigrant | 0,94 (0,212) | 0,94 (0,283) | 1,01 (0,022) | 1,32 (0,086) |
| Country of birth unknown | 1,01 (0,006) | 0,99 (0,009) | 1,02 (0,006) | 1,11 (0,031) |
| MA of Ribeirão preto | 0,85 (0,146) | 0,85 (0,018) | 0,92 (0,014) | 2,29 (0,083) |
| MA of Sorocaba | 0,70 (0,012) | 0,70 (0,012) | 0,94 (0,012) | 5,89 (0,058) |
| MA of Campinas | 0,84 (0,008) | 0,84 (0,009) | 0,96 (0,008) | 2,87 (0,051) |
| MA of Vale Paraíba | 0,84 (0,009) | 0,84 (0,012) | 0,97 (0,009) | 2,86 (0,057) |
| MA of São Paulo | 0,72 (0,005) | 0,71 (0,005) | 0,89 (0,004) | 4,29 (0,038) |
| MA of Baixada Santista | 0,75 (0,008) | 0,72 (0,008) | 0,93 (0,007) | 4,26 (0,045) |
| ACF in Institution | 0,72 (0,277) | 0,70 (0,030) | 0,99 (0,287) | 5,18 (0,206) |
| Community-based ACF | 0,72 (0,029) | 0,71 (0,031) | 0,95 (0,298) | 3,96 (0,209) |
| Contact investigation | 0,78 (0,028) | 0,80 (0,035) | 0,99 (0,028) | 2,02 (0,209) |
| PCF in Hospital | 0,67 (0,028) | 0,67 (0,028) | 0,96 (0,027) | 5,75 (0,203) |
| PCF in emergence room | 0,68 (0,027) | 0,68 (0,029) | 0,97 (0,027) | 4,96 (0,202) |
| PCF in outpatient clinics | 0,69 (0,027) | 0,69 (0,029) | 0,96 (0,027) | 5,30 (0,209) |
| Unknown case detection | 0,72 (0,029) | 0,70 (0,03) | 0,99 (0,029) | 5,30 (0,209) |
| Pulmonary TB | 1,04 (0,006) | 1,03 (0,009) | 1,00 (0,006) | 0,92 (0,027) |
| Positive microbiological status | 1,01 (0,004) | 1,02 (0,008) | 1,00 (0,005) | 0,92 (0,024) |
| Unknown microbiological status | 0,93 (0,008) | 0,78 (0,009) | 0,98 (0,007) | 1,25 (0,030) |
| Positive HIV status | 0,87 (0,008) | 0,84 (0,009) | 1,00 (0,005) | 0,87 (0,036) |
| Unknown HIV status | 0,81 (0,006) | 0,78 (0,007) | 0,99 (0,007) | 2,51 (0,025) |
| Diabetes Mellitus | 1,03 (0,008) | 1,03 (0,011) | 1,00 (0,007) | 0,87 (0,036) |
| Alcohol user | 1,01 (0,005) | 1,00 (0,008) | 0,99 (0,005) | 0,94 (0,026) |
| Smoking | 0,96 (0,005) | 0,98 (0,007) | 0,99 (0,004) | 1,03 (0,024) |
| Drug user | 0,93 (0,005) | 0,93 (0,007) | 0,98 (0,005) | 1,25 (0,026) |
| No comorbidities | 1,06 (0,004) | 1,06 (0,006) | 1,00 (0,004) | 0,78 (0,021) |
| Number of parameters | 36 | 36 | 72 | |
| log L | -277.883,8 | -254.795,2 | -217.970,1 | |
| Pseudo-R² | 2,16% | 1,1% | 51,6% | |
| AIC | 555.839,7 | 509.664,5 | 436.084,3 | |
| BIC | 555.839,7 | 510.026,2 | 436.788,0 | |

Abbreviations: PRM – Poisson regression model. NBRM – Negative binomial regression model. ZIP – Zero-Inflated Poisson. PR – prevalence ratio. OR – odds ratio. SE – standard error. TB – tuberculosis. MA – Metropolitan area. PCF – Passive case-finding. ACF – Active case finding. log L - log likelihood. Pseudo-R² - McFadden's adjusted coefficient of determination. AIC – Akaike's information criterion. BIC – Bayesian information criterion.

### Supplementary methods: Collider bias

We aimed to evaluate the characteristics of index TB cases among contacts. However, these factors may be associated with the likelihood of contacts being examined, as well as other unknown factors that could influence both this probability and TB itself. Consequently, access to examination becomes a collider, and restricting the analysis to only those contacts who were examined creates an alternative path that can lead to a spurious relationship (collider bias), as shown in the figure below. Therefore, by weighting by the inverse probability of being examined among those screened, we avoid this alternative path and thus minimise the introduction of bias in the association between risk factors and TB.

**Risk factors for TB**

**Tuberculosis**

**Other risk factors for TB**

**Examined contacts**

**Note:**

Exposure	Outcome	Unobserved variable	Adjusted variable

Causal path	Biasing path

**Figure S5.** Representation of collider bias when restricting the analysis to examined contacts in the study.

### Supplementary methods: Hierarchical conceptual framework

| Distal variables |  | -Temporal: year of diagnosis  -Geographic characteristics: metropolitan area of residence |
| --- | --- | --- |
| Intermediate variables I |  | -Sociodemographic characteristics: sex, age group, race or ethnicity and county of birth |
| Intermediate variables II |  | -Socioeconomic characteristics: education, homeless and incarcerated;  -Health characteristics: HIV status and comorbidities |
| Proximal variables |  | -Case detection strategies;  -Clinical characteristics: chest X-ray, anatomical classification, microbiological status and drug-resistant |
| **Active TB among contacts** |  |  |

**Figure S6.** Hierarchical conceptual framework for adjusted multilevel Poisson regression models.

### Supplementary results: Effect of municipalities on TB prevalence

**Table S3.** Epidemiological indicators, predict variation, and performance of tuberculosis contact investigation strategy by municipality in the state of São Paulo, Brazil, from 2010 to 2020.

| **Municipality** | **New cases**  **of TB** | **Incidence rate** | **Multilevel Poisson regression model** | | | **Contacts** | | | **Yield of TB** |
| --- | --- | --- | --- | --- | --- | --- | --- | --- | --- |
|  | **N** | **New cases/**  **100,000 PPY** | **Predict**  **Value** | **Lower**  **95% CI** | **Upper**  **95% CI** | **Screened** | **Examined** | **Active TB** | **%** |
|  |  |  |  |  |  | **N** | **N** | **N** |  |
| **Above average (n=43)** |  |  |  |  |  |  |  |  |  |
| Ibiúna | 158 | 17,8 | 1,44 | 1,14 | 1,75 | 483 | 307 | 23 | 4,8 |
| Ituverava | 92 | 18,2 | 1,13 | 0,59 | 1,66 | 217 | 189 | 23 | 10,6 |
| Piracaia | 63 | 19,1 | 1,08 | 0,45 | 1,71 | 185 | 60 | 9 | 4,9 |
| Itapuí | 32 | 21,2 | 1,03 | 0,46 | 1,59 | 166 | 160 | 16 | 9,6 |
| Itaguaí | 27 | 19,3 | 1,01 | 0,30 | 1,72 | 71 | 69 | 11 | 15,5 |
| Campos do Jordao | 68 | 10,8 | 1,00 | 0,45 | 1,56 | 192 | 197 | 21 | 10,9 |
| Bady Bassitt | 43 | 21,5 | 0,97 | 0,45 | 1,49 | 139 | 129 | 18 | 12,9 |
| Ribeirão Preto | 2134 | 26,4 | 0,88 | 0,74 | 1,03 | 4271 | 3048 | 243 | 5,7 |
| Botucatu | 338 | 20,1 | 0,88 | 0,57 | 1,18 | 1188 | 1157 | 47 | 4,0 |
| Iaras | 92 | 88,1 | 0,84 | 0,03 | 1,64 | 90 | 85 | 7 | 7,8 |
| Jau | 320 | 18,7 | 0,82 | 0,38 | 1,26 | 850 | 646 | 29 | 3,4 |
| Iperó | 315 | 76,1 | 0,80 | 0,26 | 1,35 | 298 | 233 | 14 | 4,7 |
| Sarapuí | 10 | 9,2 | 0,78 | 0,09 | 1,46 | 24 | 17 | 7 | 29,2 |
| Jaci | 22 | 26,0 | 0,76 | 0,01 | 1,50 | 915 | 906 | 27 | 3,0 |
| Votuporanga | 188 | 15,9 | 0,73 | 0,28 | 1,18 | 468 | 423 | 26 | 5,6 |
| Iguape | 177 | 54,6 | 0,71 | 0,29 | 1,14 | 510 | 421 | 48 | 9,4 |
| Itapira | 187 | 20,7 | 0,71 | 0,22 | 1,20 | 423 | 379 | 31 | 7,3 |
| Itatiba | 240 | 17,6 | 0,71 | 0,35 | 1,06 | 956 | 687 | 33 | 3,5 |
| Atibaia | 354 | 21,7 | 0,69 | 0,18 | 1,20 | 441 | 244 | 37 | 8,4 |
| Laranjal Paulista | 115 | 32,9 | 0,69 | 0,27 | 1,10 | 328 | 317 | 22 | 6,7 |
| Indaiatuba | 480 | 17,4 | 0,69 | 0,38 | 0,99 | 1053 | 724 | 53 | 5,0 |
| Capivari | 146 | 23,7 | 0,67 | 0,18 | 1,16 | 409 | 331 | 22 | 5,4 |
| Cachoeira Paulista | 64 | 16,1 | 0,66 | 0,10 | 1,22 | 169 | 131 | 26 | 15,4 |
| Pontal | 150 | 29,2 | 0,65 | 0,14 | 1,16 | 471 | 378 | 14 | 3,0 |
| São Paulo | 66788 | 47,3 | 0,64 | 0,62 | 0,67 | 166251 | 95902 | 5417 | 3,3 |
| Presidente Prudente | 771 | 28,0 | 0,61 | 0,23 | 0,99 | 727 | 642 | 31 | 4,3 |
| Cruzeiro | 337 | 33,8 | 0,60 | 0,26 | 0,94 | 1082 | 725 | 32 | 3,0 |
| Sertãozinho | 365 | 25,1 | 0,58 | 0,19 | 0,98 | 715 | 441 | 21 | 2,9 |
| Várzea Paulista | 327 | 23,3 | 0,55 | 0,14 | 0,96 | 1018 | 594 | 35 | 3,4 |
| Santa Isabel | 221 | 34,6 | 0,54 | 0,10 | 0,97 | 622 | 452 | 26 | 4,2 |
| Registro | 345 | 52,7 | 0,53 | 0,14 | 0,92 | 989 | 786 | 31 | 3,1 |
| Itapevi | 1196 | 47,1 | 0,52 | 0,37 | 0,67 | 4337 | 3336 | 254 | 5,9 |
| Sorocaba | 2208 | 28,2 | 0,51 | 0,29 | 0,73 | 3427 | 1023 | 63 | 1,8 |
| Bertioga | 410 | 65,4 | 0,47 | 0,18 | 0,76 | 867 | 800 | 60 | 6,9 |
| Santana de Parnaíba | 387 | 25,5 | 0,47 | 0,11 | 0,83 | 996 | 734 | 30 | 3,0 |
| Assis | 335 | 27,9 | 0,45 | 0,00 | 0,91 | 806 | 698 | 19 | 2,4 |
| Guarulhos | 5543 | 36,0 | 0,44 | 0,34 | 0,53 | 15541 | 9425 | 403 | 2,6 |
| Bauru | 1995 | 46,2 | 0,43 | 0,16 | 0,71 | 2431 | 1713 | 55 | 2,3 |
| Araras | 342 | 21,8 | 0,43 | 0,00 | 0,85 | 823 | 701 | 21 | 2,6 |
| Jundiai | 1101 | 22,7 | 0,35 | 0,13 | 0,58 | 2305 | 1990 | 78 | 3,4 |
| Campinas | 3779 | 27,1 | 0,35 | 0,22 | 0,47 | 11413 | 7808 | 317 | 2,8 |
| Embu das Artes | 1270 | 42,8 | 0,31 | 0,14 | 0,48 | 4398 | 4010 | 122 | 2,8 |
| Piracicaba | 1802 | 38,5 | 0,26 | 0,01 | 0,50 | 2677 | 2216 | 56 | 2,1 |
| **Below average (n=21)** |  |  |  |  |  |  |  |  |  |
| Suzano | 1100 | 32,7 | -1,05 | -1,46 | -0,64 | 2970 | 2420 | 31 | 1,0 |
| Santa Barbara D'oeste | 389 | 16,7 | -1,00 | -1,66 | -0,35 | 1039 | 972 | 2 | 0,2 |
| São Caetano do Sul | 292 | 14,2 | -0,95 | -1,66 | -0,24 | 555 | 444 | 1 | 0,2 |
| Mairinque | 154 | 28,6 | -0,92 | -1,71 | -0,13 | 498 | 395 | 0 | 0,0 |
| Pindamonhangaba | 355 | 18,5 | -0,88 | -1,49 | -0,27 | 1014 | 778 | 4 | 0,4 |
| Tupã | 180 | 21,1 | -0,86 | -1,66 | -0,06 | 463 | 448 | 0 | 0,0 |
| Itapetininga | 644 | 35,1 | -0,83 | -1,41 | -0,26 | 1125 | 822 | 11 | 1,0 |
| Araraquara | 469 | 16,5 | -0,82 | -1,47 | -0,17 | 946 | 661 | 4 | 0,4 |
| São Jose dos Campos | 1853 | 22,4 | -0,70 | -1,08 | -0,31 | 3883 | 1922 | 22 | 0,6 |
| Sumaré | 697 | 21,9 | -0,63 | -1,05 | -0,20 | 1860 | 1175 | 22 | 1,2 |
| São Vicente | 4636 | 113,9 | -0,62 | -0,80 | -0,44 | 9517 | 7152 | 92 | 1,0 |
| Guaratinguetá | 336 | 23,1 | -0,59 | -1,13 | -0,06 | 909 | 728 | 11 | 1,2 |
| Lorena | 300 | 28,9 | -0,59 | -1,14 | -0,04 | 954 | 686 | 6 | 0,6 |
| Itu | 475 | 23,9 | -0,55 | -1,03 | -0,07 | 1015 | 713 | 12 | 1,2 |
| Cotia | 823 | 30,3 | -0,53 | -0,89 | -0,17 | 2283 | 2111 | 35 | 1,5 |
| Itapecerica da Serra | 800 | 41,2 | -0,42 | -0,81 | -0,03 | 1706 | 1329 | 24 | 1,4 |
| Cubatão | 1176 | 79,8 | -0,41 | -0,66 | -0,16 | 3049 | 2535 | 52 | 1,7 |
| Hortolândia | 1087 | 41,2 | -0,37 | -0,73 | -0,01 | 1738 | 1458 | 30 | 1,7 |
| Mauá | 1647 | 30,5 | -0,32 | -0,63 | -0,01 | 3592 | 1944 | 28 | 0,8 |
| Praia Grande | 2954 | 86,6 | -0,29 | -0,48 | -0,10 | 6736 | 3949 | 88 | 1,3 |
| Guarujá | 2967 | 82,3 | -0,27 | -0,44 | -0,10 | 7161 | 4432 | 89 | 1,2 |

*Note:* Age-standardized incidence rate using WHO standard population [18].

Abbreviations: TB – Tuberculosis. PPY – Population per year. 95% CI – 95% confidence interval.
